# Puberty differentially predicts brain maturation in male and female youth: A longitudinal ABCD Study

**DOI:** 10.1101/2022.12.22.22283852

**Authors:** Dani Beck, Lia Ferschmann, Niamh MacSweeney, Linn B. Norbom, Thea Wiker, Eira Aksnes, Valerie Karl, Fanny Dégeilh, Madelene Holm, Kathryn L. Mills, Ole A. Andreassen, Ingrid Agartz, Lars T. Westlye, Tilmann von Soest, Christian K. Tamnes

## Abstract

Research has demonstrated associations between pubertal development and brain maturation. However, existing studies have been limited by small samples, cross-sectional designs, and inconclusive findings regarding directionality of effects and sex differences.

We examined the longitudinal temporal coupling of puberty status assessed using the Pubertal Development Scale (PDS) and magnetic resonance imaging (MRI)-based grey and white matter brain structure. Our sample consisted of 8,896 children and adolescents at baseline (mean age = 9.9) and 6,099 at follow-up (mean age = 11.9) from the Adolescent Brain and Cognitive Development (ABCD) Study.

Applying multigroup Bivariate Latent Change Score (BLCS) models, we found that baseline PDS predicted the rate of change in cortical thickness among females and rate of change in cortical surface area for both males and females. We also found a correlation between baseline PDS and surface area and co-occurring changes over time in males. Diffusion tensor imaging (DTI) analysis revealed correlated change between PDS and fractional anisotropy (FA) for both males and females, but no significant associations for mean diffusivity (MD).

Our results suggest that pubertal status predicts cortical maturation, and that the strength of the associations differ between sex. Further research is needed to understand the impact of environmental and lifestyle factors.

## 1. Introduction

Adolescence is a transitional stage from childhood to adulthood that is marked by prolonged tissue-specific neurodevelopmental processes. Tethered to this, puberty has been highlighted as a defining period for protracted brain structural maturation during adolescence, with a range of studies linking concurrent trends of pubertal and brain development that go beyond the effects of age (Ando et al., 2021; Blakemore et al., 2010; Herting & Sowell, 2017; Vijayakumar et al., 2021). However, existing studies on puberty-brain associations are limited by small sample sizes and cross-sectional designs, in addition limited research on directionality of effects, underscoring the need for large-scale longitudinal studies that examine the temporal coupling of pubertal development and brain maturational processes.

Although pubertal development is characterised by progress through the same stages for each individual — involving changes in secondary sex characteristics, rising hormone levels, and emerging reproductive capability — there is a large variation in the timing and tempo of these changes. The variation in pubertal timing, defined as an individual’s relative pubertal developmental stage compared with same -age and -sex peers (Ullsperger & Nikolas, 2017), involve both inter-individual and between-sex differences, with the typical age of pubertal onset ranging from 9 to 14 years in males and 8 to 13 years in females (Herting et al., 2021; Savin-Williams & Ream, 2006).

Additionally, there is a genetic contribution, with heritability estimates ranging from 50 to 80% (Stroud & Davila, 2011). A growing body of evidence has also linked variation in pubertal timing to lifestyle and environmental factors, including childhood obesity (Ahmed et al., 2009; Herting et al., 2021) and socioeconomic status (SES) (Braithwaite et al., 2009; Herting et al., 2021). An implication of this is that pubertal timing may be partially malleable and, as such, represents a window of opportunity for early intervention that targets youth at risk of health issues related to pubertal timing. For example, earlier pubertal onset is associated with an increased risk of mental health outcomes related to risk-taking (Braams et al., 2015; Collado-Rodriguez et al., 2014), depression and anxiety (Copeland et al., 2019; MacSweeney et al., 2023; McNeilly et al., 2022; Mendle et al., 2010; Mendle & Ferrero, 2012), and substance use (Marceau et al., 2019; Patton et al., 2004; Stumper et al., 2019).

In conjunction to pubertal development, the brain shows continued tissue-specific changes that are prone to influence by fluctuating environmental pressures (Ferschmann et al., 2022; Tooley et al., 2021) beyond the complex genetic contribution (Grasby et al., 2020). Further, puberty has been implicated as an influencing factor to the rate of maturation in the brain, and as such recommended as non-negligible when studying adolescent brain maturation (Holm et al., 2022; Wierenga et al., 2018).

Previous cross-sectional (Bramen et al., 2011; Paus et al., 2010; Pfefferbaum et al., 2016) and longitudinal (Herting et al., 2015; Nguyen et al., 2013; Vijayakumar et al., 2021) studies have reported reductions in global grey matter volume and cortical thickness associated with higher pubertal status and testosterone levels, mirroring normative developmental patterns of grey matter during adolescence (Mills et al., 2016; Tamnes et al., 2017). However, when controlling for age, findings have been less consistent, with some studies reporting associations between decreased cortical grey matter and more pubertally developed females (Bramen et al., 2011; Peper, Brouwer, et al., 2009), others reporting positive associations between grey matter volume and testosterone in males (Peper, Brouwer, et al., 2009), and several reporting null findings for both sexes (Bramen et al., 2011; Koolschijn et al., 2014; Peper, Brouwer, et al., 2009; Peper, Schnack, et al., 2009).

For white matter, numerous cross-sectional studies have reported positive associations between pubertal status and white matter volumes (Chavarria et al., 2014; Perrin et al., 2009; Pfefferbaum et al., 2016), with some conflicting findings (Peper, Schnack, et al., 2009). Similarly, studies have reported positive associations between testosterone levels and white matter volume among males (Paus et al., 2010; Perrin et al., 2008), with inconsistent findings when accounting for age (Peper, Brouwer, et al., 2009).

Diffusion tensor imaging (DTI) provides information regarding white matter architecture and microstructure. DTI studies have generally reported higher fractional anisotropy (FA) and lower mean diffusivity (MD) among more pubertally developed adolescents (Herting et al., 2012; Menzies et al., 2014), mirroring normative developmental patterns (Lebel & Deoni, 2018; Pfefferbaum et al., 2016). However, to date, only one longitudinal study (Herting et al., 2017) has investigated associations between puberty and white matter microstructure using DTI, albeit with a small sample, finding that physical pubertal changes predicted changes in white matter. However, further longitudinal research is necessary to validate and extend previous findings and to examine potential sex differences.

Using a large-scale longitudinal sample (N = 8,896 at baseline; N = 6,099 at follow-up) of children and adolescents between the ages of 9-14 years from the Adolescent Brain Cognitive Development (ABCD) Study cohort, we investigated the developmental associations between puberty and brain structure and tested whether and to what extent these associations differ between male and female youth. By leveraging a powerful Structural Equation Modeling (SEM) framework, multigroup Bivariate Latent Change Scores (BLCS), we examined how individual and sex differences in pubertal development relate to differences in structural brain maturation. Importantly, BLSC allows us to test the temporal dynamics of the puberty-brain relationship, specifically whether pubertal status and brain features correlate at baseline, brain structure at baseline predicts change in pubertal status, baseline pubertal status predicts change in brain structure, and whether changes in puberty and brain structure co-occur over the course of the study period.

Based on the most consistently reported effects documented to date, we expected cross-sectional and longitudinal associations between pubertal status measured with the Pubertal Development Scale (PDS) and global features of cortical thickness and surface area and white matter microstructure (FA, MD) in the examined age range. Specifically, based on morphometrical and DTI studies (Bramen et al., 2011; Herting et al., 2012, 2015; Menzies et al., 2014; Nguyen et al., 2013; Paus et al., 2010; Pfefferbaum et al., 2016; Vijayakumar et al., 2021) reporting effects that mirror normative developmental patterns, we hypothesised correlations at baseline across all brain MRI features and for changes in pubertal status and brain structure to co-occur. In terms of coupling effects (i.e., regressions), despite limited research on directionality of effects, we hypothesised that baseline pubertal status would predict change in global brain structure and not vice versa, in line with a previous study reporting pubertal changes predicting changes in brain maturation (Herting et al., 2017). Additionally, based on previous research reporting more consistent effects for females compared to males, we hypothesised sex related effects to be present, with generally stronger puberty-brain associations in females compared to males.

## 2. Methodology

### 2.1. Description of sample

The initial sample consisted of children and adolescents that are part of the ongoing longitudinal ABCD Study, including a baseline cohort of ∼11,800 nine-and ten-year-olds to be followed for the course of ten years (Garavan et al., 2018). Data used in the present study were drawn from the ABCD curated annual release 4.0, containing data from baseline up until the second-year visit (https://data-archive.nimh.nih.gov/abcd). All ABCD Study data is stored in the NIMH Data Archive Collection #2573, which is available for registered and authorised users (Request #7474, PI: Westlye). The 4.0 release will be permanently available as a persistent dataset defined in the NDA Study 1299 and has been assigned the DOI 10.15154/1523041.

To minimize confounding effects from complex family-related factors, unrelated participants were chosen by randomly selecting an individual from each family ID, subsequently excluding any siblings (see Supporting Information (SI) Section 1 for details). Additionally, participants were excluded using the ABCD Study exclusion criteria, which included non-English proficiency, general MRI contraindications, a history of a major neurological disorder, traumatic brain injury, extreme premature birth (<28 weeks gestational age), a diagnosis of schizophrenia, intellectual disability, moderate to severe autism spectrum disorder, or substance abuse disorder (Karcher et al., 2018).

Following procedures of quality control (see SI Section 1) for each brain measure (FA, MD, cortical thickness, surface area), PDS, and our covariates of interest including SES, body-mass index (BMI), and genetic ancestry-derived ethnicity, the final sample consisted of N = 8,896 participants at baseline (4,223 females, 4,673 males) and N = 6,099 participants at follow-up (2,798 females, 3,301 males). Attrition analyses can be found in SI Section 1 and SI Figure 1. Table 1 displays means and standard deviations of each brain MRI metric, PDS, and covariates of interest, split by sex, while the age and PDS distributions in the sample are presented in Figure 1. Ethical approvals for the study are described in SI Section 2.

**Figure 1.**
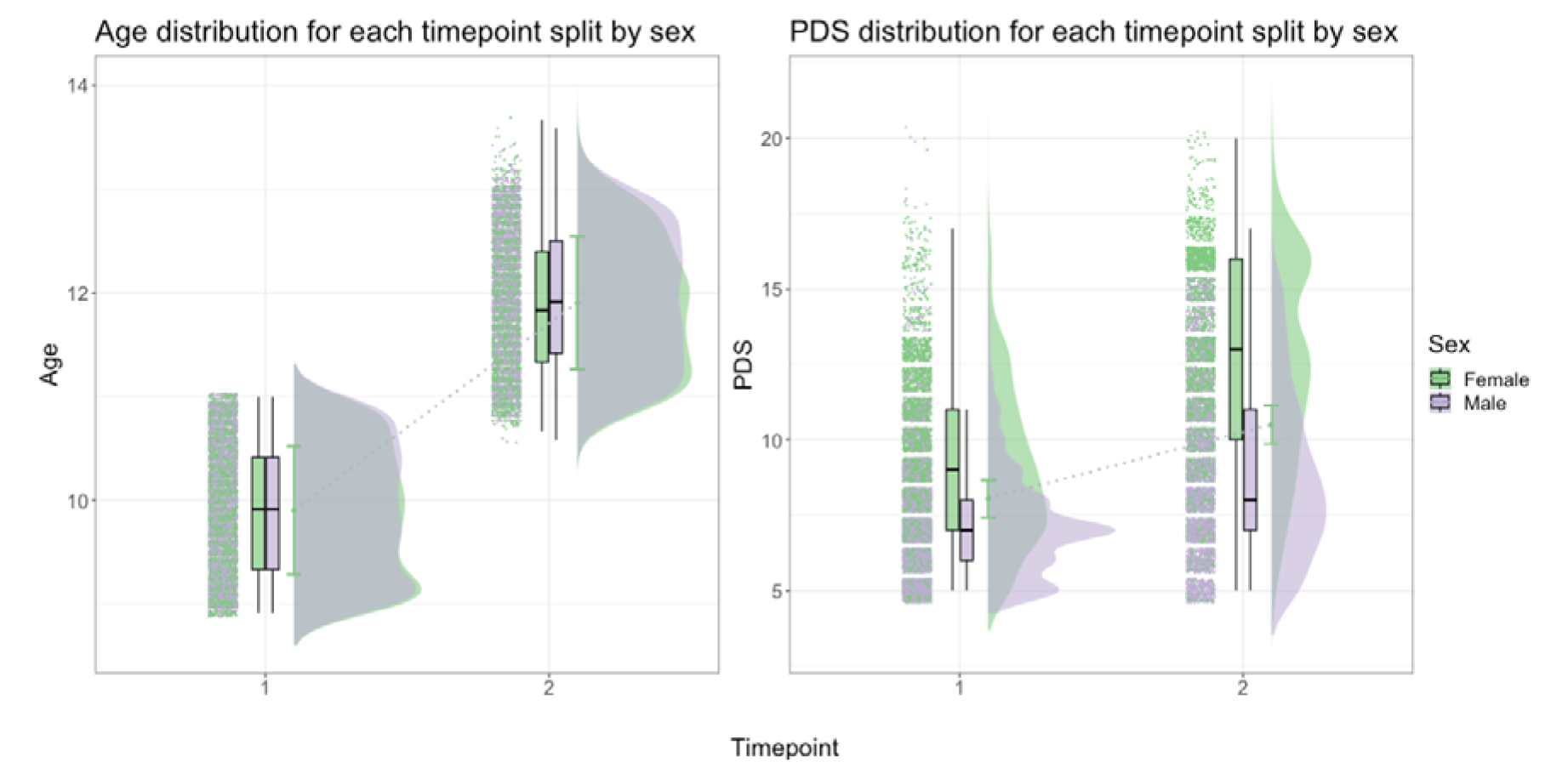
Age (left) and PDS (right) distribution of the study sample, split by sex and timepoint. Dotted line between plots connects the mean age and PDS values at baseline (timepoint 1) and follow-up (timepoint 2). Age and total PDS scores shown as distributions where green represents females and purple represents males.

**Table 1.** Descriptive statistics of the sample, including mean ± standard deviation (SD) for each measure.

|  | **Baseline** | **Follow-up** | **Males at** | **Females at** | **Males at** | **Females at** |
| --- | --- | --- | --- | --- | --- | --- |
|  | **sample** | **sample** | **baseline** | **baseline** | **follow-up** | **follow-up** |
| **N subjects** | 8,896 | 6,099 | 4,673 | 4,223 | 3,301 | 2,798 |
| **Age** | 9.9 ± 0.62 | 11.9 ± 0.64 | 9.9 ± 0.62 | 9.9 ± 0.61 | 11.9 ± 0.64 | 11.9 ± 0.63 |
| **PDS** | 8.0 ± 2.40 | 10.5 ± 3.48 | 7.2 ± 1.86 | 8.9 ± 2.59 | 8.9 ± 2.74 | 12.4 ± 3.30 |
| **FA** | 0.503 ± 0.02 | 0.509 ± 0.02 | 0.503 ± 0.02 | 0.503 ± 0.02 | 0.509 ± 0.02 | 0.510 ± 0.02 |
| **MD** | 0.800 ± 0.02 | 0.789 ± 0.02 | 0.802 ± 0.03 | 0.790 ± 0.02 | 0.792 ± 0.02 | 0.786 ± 0.02 |
| **CT** | 2.72 ± 0.09 | 2.69 ± 0.09 | 2.72 ± 0.09 | 2.73 ± 0.09 | 2.69 ± 0.09 | 2.69 ± 0.09 |
| **SA** | 1,899 ± 180 | 1,907 ± 183 | 1,977 ± 167 | 1,813 ± 154 | 1,990 ± 168 | 1,814 ± 153 |
| **BMI** | 18.82 ± 4.13 | 20.57 ± 4.84 | 18.75 ± 4.03 | 18.91 ± 4.23 | 20.35 ± 4.71 | 20.84 ± 4.98 |
| **SES** | 7.25 ± 2.39 | 7.56 ± 2.23 | 7.26 ± 2.39 | 7.24 ± 2.39 | 7.58 ± 2.44 | 7.53 ± 2.21 |

**Note:** MRI metrics of FA, MD, cortical thickness (CT), and surface area (SA) represent scanner-harmonised scores. CT represented in mm. Area represented in cm<sup>2</sup>. SES represents a total score derived from the average income from both parental and partner income, where 7 represents those earning between \$50-75k annually.

### 2.2. MRI acquisition and processing

Neuroimaging data were acquired at 21 different sites and processed by the ABCD team. A 3-T Siemens Prisma, General Electric 750 or Phillips scanner was used for data acquisition. Protocols used for data acquisition and processing are described elsewhere (Casey et al., 2018; Hagler et al., 2019). In brief, T1-weighted data was acquired by magnetisation-prepared rapid acquisition gradient echo scans with a resolution of 1×1×1 mm^3^, which was used for generating cortical structural measures, and diffusion-weighted data was obtained by high angular resolution diffusion imaging scans, used for generating white matter microstructural measures.

Two modalities of brain structural measures were used in the present study: grey matter cortical measures and white matter microstructural measures (Hagler et al., 2019). Cortical reconstruction and volumetric segmentation was performed with FreeSurfer 6.0 (Dale et al., 1999; Fischl et al., 2002). White matter microstructural measures (DTI) were generated using AtlasTrack, a probabilistic atlas-based method for automated segmentation of white matter fiber tracts (Hagler Jr. et al., 2009). For cortical measures, global measures were generated for cortical surface area and thickness. For DTI, measures of FA and MD were generated over the whole brain. For full pipeline, see SI Section 3.

### 2.3. Multi-site MRI scanner effects

To adjust for non-biological variance introduced by multi-scanner MRI and acquisition protocols (i.e., scanner effects), a longitudinal harmonisation technique was carried out using the R package *LongComBat* (Beer et al., 2020). To reduce bias in scaling and spatial resolution of features, our four global brain MRI measures (FA, MD, cortical thickness, surface area) were harmonised separately. Since the harmonisation is applied to model residuals, the longitudinal ComBat model should ideally match the model in the final analysis. As such, each model was fitted with formulas including age, PDS, BMI, SES, and genetic ancestry-derived ethnicity as covariates. SI Figures 2 and 3 show distribution of each MRI measure before harmonisation and after longitudinal ComBat.

**Figure 2.**
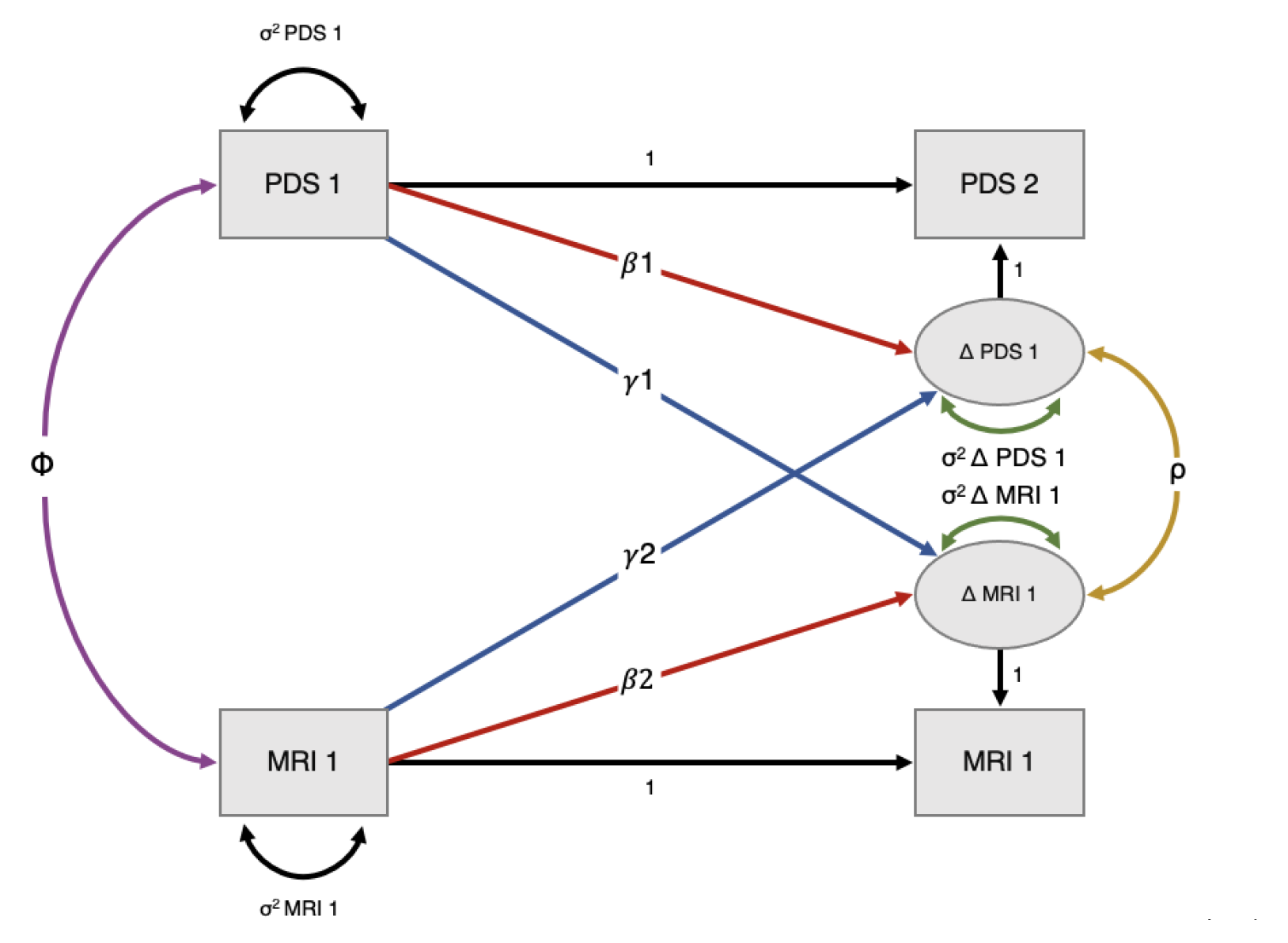
Bivariate latent change score (BLCS) model with two timepoints, examining correlation between pubertal status (PDS) and brain MRI metric (MRI) at baseline (pink), to what extent pubertal status at baseline predicts the rate of change in brain MRI metric (blue: 1), to what extent brain MRI metric at baseline predicts the rate of change in pubertal status (blue: 2), and whether changes in PDS and MRI co-occur (yellow) after considering the coupling (1, 2) pathways. Image adapted from Kievit et al. (2018) with permission from author.

**Figure 3.**
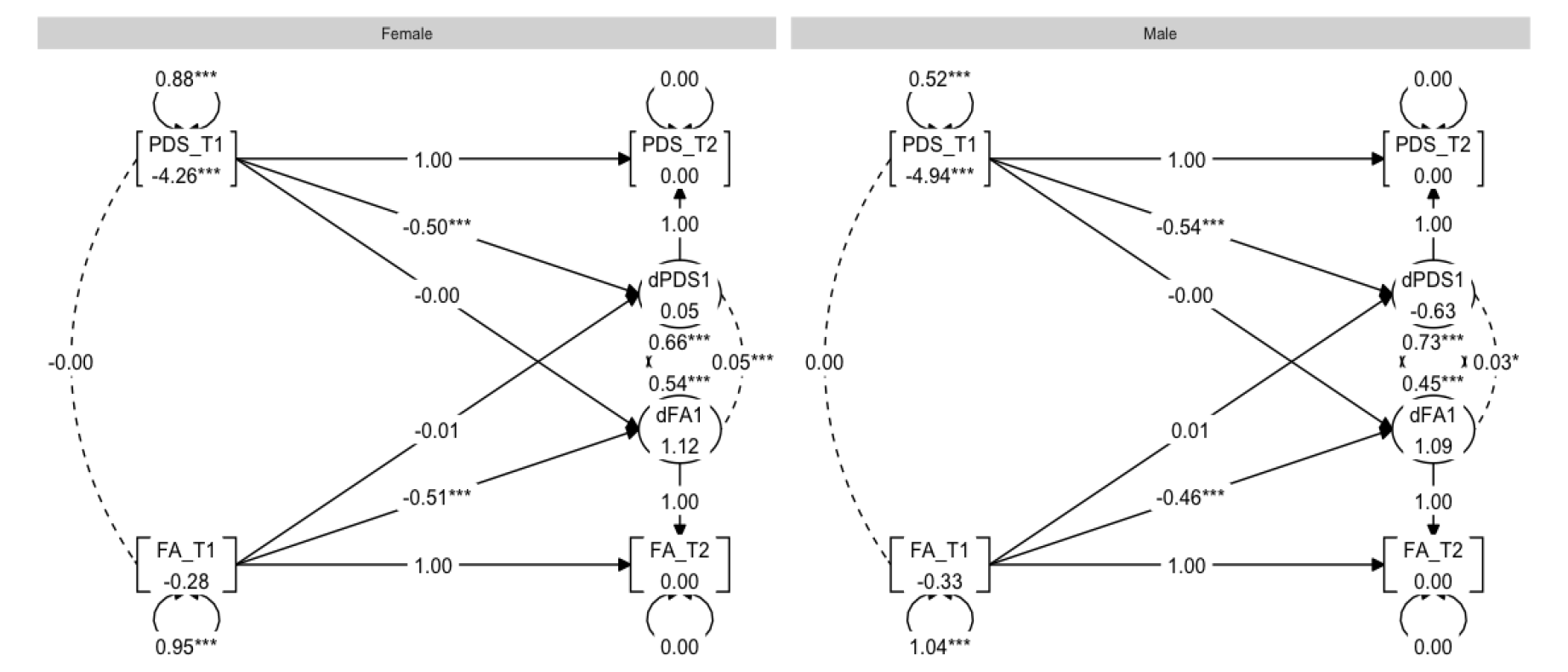
Graph showing the multigroup BLCS models for fractional anisotropy (FA) and PDS relations.

### 2.4. Pubertal development assessment

Pubertal status was measured using the PDS (Petersen et al., 1988) to examine the perceived development of secondary sex characteristics such as growth spurts, body hair growth, skin changes, breast development and menarche in females, and voice changes and growth of testes in males. The PDS is a questionnaire designed to mimic the traditional Tanner staging assessment without the use of reference pictures (Petersen et al., 1988). Due to previous research showing that youth tend to over-report their perceived physical development at younger ages (Schlossberger et al., 1992), the current study utilised caregiver PDS report.

The PDS includes five-items, each rated on a 4-point scale (1 = no development; 2 = development has barely begun; 3 = development is definitely underway; and 4 = development is complete; except menstruation, which is coded 1 = has not begun, 4 = has begun). Thus, higher scores reflect more advanced pubertal development. The PDS has shown high inter-rater reliability between parent and self-rated assessment to clinicians, and correlates highly with plasma levels of gonadal hormones (Carskadon & Acebo, 1993; Koopman-Verhoeff et al., 2020) in addition to the Tanner stages (Koopman-Verhoeff et al., 2020). We used total PDS score for our analyses. When describing our results, PDS and the term *pubertal status* is used interchangeably for cross-sectional analyses, while *pubertal development* is used for longitudinal analyses. Note, both pubertal status and development refer to individual differences in PDS scores while controlling for age.

### 2.5. Covariates of interest

Previous research has highlighted several factors that potentially contribute to pubertal timing and development, including BMI, SES, and ethnicity (Ahmed et al., 2009; Braithwaite et al., 2009; Deardorff et al., 2019; Freedman et al., 2002; Herting et al., 2021; Kaplowitz et al., 2001). Therefore, we included BMI, SES, and genetic ancestry-derived ethnicity, in addition to age, as covariates. For full details on how each covariate was measured and derived, see SI Section 4. For figures relating to distribution, frequency, and associations between covariates, see SI Figures 4-6.

**Figure 4.**
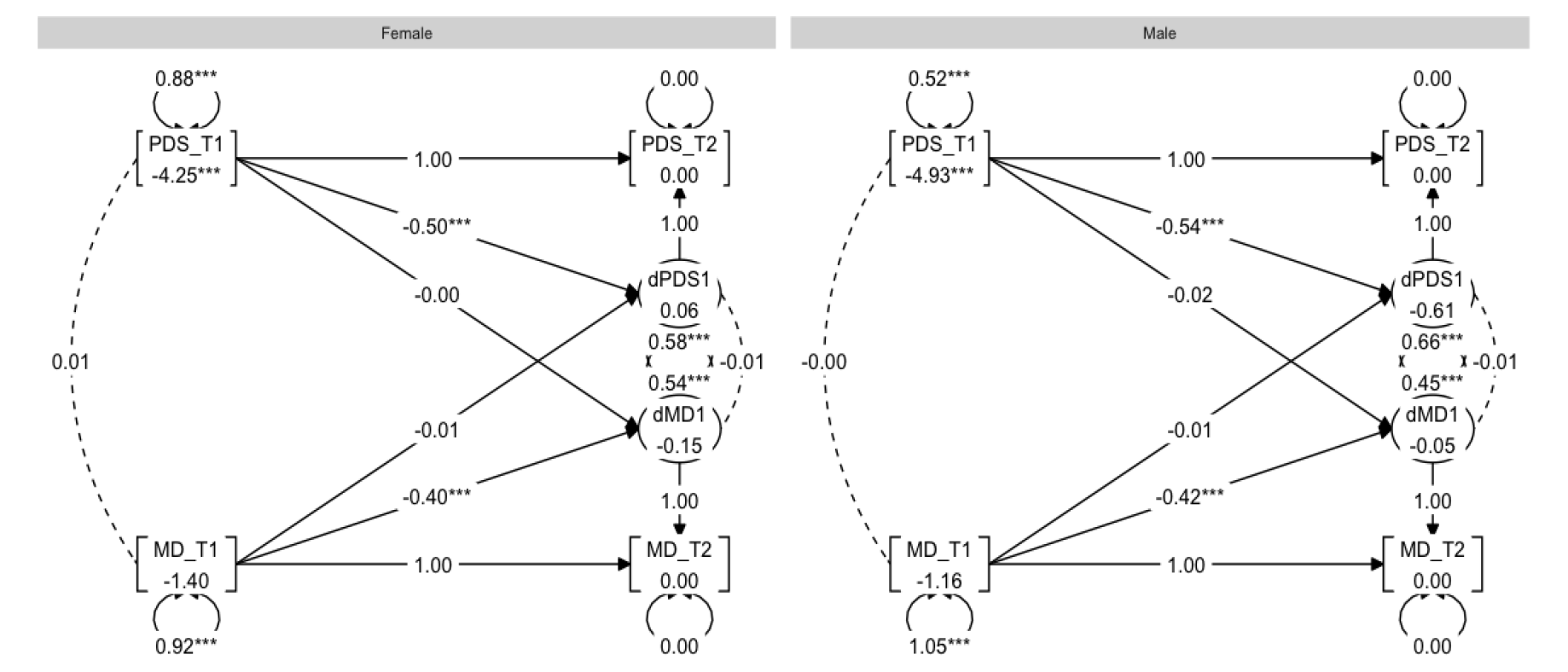
Graph showing the multigroup BLCS models for mean diffusivity (MD) and PDS relations.

### 2.6. Statistical analysis

The code used in the present study is available in a public repository - Open Science Framework (OSF) - and accessible directly through the OSF webpage https://osf.io/nh649/.

All analyses were carried out using R version 4.0.0 (R Core Team, 2022). A multigroup BLCS model was tested using the *lavaan* package (Rosseel, 2012) following recommendations provided by Kievit et al. (2018) for model setup.

The rationale behind using BLCS models is rooted in its ability to model latent change by using only two data collection waves (Kievit et al., 2018). Moreover, the modelling framework is especially powerful for testing cross-domain couplings which captures the extent to which change in one domain is a function of the starting level in the other. Further motivation for using this model was our interest in sex-related effects. Using a multi-group BLCS model allows one to assess four different brain-puberty associations of interest for each sex in one model, and to test whether the size of associations significantly differ across sex.

Four separate models were set up to examine 1) correlations at baseline between pubertal status and MRI measures of global FA, MD, cortical thickness, and surface area; 2) to what extent the baseline pubertal status predicted rate of change in global brain MRI measures; 3) to what extent the baseline score of global brain MRI measure predicted rate of change in the pubertal development; and lastly, 4) whether change co-occurred. To investigate sex differences in the associations, we used multiple group analyses in the framework of SEM, with sex (female, male) as the grouping variable.

We quantified to what extent change in pubertal status (ΔPDS) between baseline and follow-up was a function of brain MRI measure (*γ*2) and pubertal status (*β*_1_) at baseline as following:

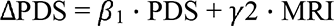

And to what extent change in brain MRI measure (MRI) between baseline and follow-up was a function of pubertal status (*γ*1) and brain MRI measure (*β*_2_) at baseline as following:

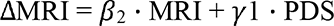

For an illustrative description of this model, see Figure 2. Due to their relatedness to pubertal development, age, BMI, SES, and genetic ancestry-derived ethnicity were included in each model. Full information for maximum likelihood estimation procedures were used for all BLCS analyses, thereby providing missing data routines that are considered to be adequate (Enders, 2010).

Analyses to test sex differences were also carried out, where the model parameters above (1-4) were constrained to be equal across the two groups and tested by means of χ^2^-difference tests against their respective non-constrained (original) model for each of the four brain MRI metrics. To test sex differences for all associations for all four models, sixteen such tests were carried. The significance threshold was set at *p* < 0.05, and the results were corrected for multiple comparisons using the false discovery rate (FDR) adjustment (Benjamini & Hochberg, 1995). Both uncorrected (*p*) and corrected (*p^corr^*) values are reported for these tests.

## 3. Results

### 3.1. Descriptive statistics

Descriptive statistics can be found in Table 1. Figures for each brain MRI measure as a function of age are presented in SI Figure 7. Brain MRI trajectories as a function of PDS are presented in SI Figure 8.

### 3.2. Multigroup bivariate latent change score (BLCS) models

The main results of the models are summarised in Table 2. For the full models, including analyses testing for sex differences in the associations, see SI Section 5.

**Table 2.** Multigroup BLCS model for each brain MRI metric, where numbers 1-4 represent the following effects: (1) correlation at baseline, (2) regression (PDS at baseline related to change in MRI metric), (3) regression (MRI metric at baseline related to change in PDS), (4) correlation of change score.

| Effect |  | Female |  |  |  | Male |  |  |  |
| --- | --- | --- | --- | --- | --- | --- | --- | --- | --- |
|  |  | Est | SE | z | p | Est | SE | z | p |
| **FA** |  |  |  |  |  |  |  |  |  |
| (1) | $PDS_{t1} \leftrightarrow FA_{t1} (\rho)$ | -0.00 | 0.02 | -0.27 | 0.79 | 0.00 | 0.01 | 0.14 | 0.89 |
| (2) | $PDS_{t1} \rightarrow \Delta FA (\gamma_1)$ | -0.00 | 0.02 | -0.07 | 0.95 | -0.00 | 0.03 | -0.00 | 0.99 |
| (3) | $FA_{t1} \rightarrow \Delta PDS (\gamma_2)$ | -0.01 | 0.02 | -0.83 | 0.41 | 0.01 | 0.01 | 0.50 | 0.62 |
| (4) | $\Delta PDS \leftrightarrow \Delta FA (\rho)$ | 0.05 | 0.01 | 3.54 | **<0.01** | 0.03 | 0.01 | 2.36 | **0.02** |
| **MD** |  |  |  |  |  |  |  |  |  |
| (1) | $PDS_{t1} \leftrightarrow MD_{t1} (\rho)$ | 0.01 | 0.02 | 0.53 | 0.60 | -0.00 | 0.01 | -0.30 | 0.76 |
| (2) | $PDS_{t1} \rightarrow \Delta MD (\gamma_1)$ | -0.01 | 0.02 | -0.27 | 0.79 | -0.02 | 0.03 | -0.77 | 0.44 |
| (3) | $MD_{t1} \rightarrow \Delta PDS (\gamma_2)$ | -0.01 | 0.02 | -0.59 | 0.56 | -0.01 | 0.01 | -1.03 | 0.30 |
| (4) | $\Delta PDS \leftrightarrow \Delta MD (\rho)$ | -0.01 | 0.02 | -0.59 | 0.56 | -0.01 | 0.02 | -0.55 | 0.58 |
| **CT** |  |  |  |  |  |  |  |  |  |
| (1) | $PDS_{t1} \leftrightarrow CT_{t1} (\rho)$ | -0.01 | 0.02 | -0.71 | 0.48 | 0.00 | 0.01 | 0.05 | 0.96 |
| (2) | $PDS_{t1} \rightarrow \Delta CT (\gamma_1)$ | -0.08 | 0.02 | -4.56 | **<0.01** | -0.00 | 0.02 | -0.10 | 0.92 |
| (3) | $CT_{t1} \rightarrow \Delta PDS (\gamma_2)$ | 0.01 | 0.02 | 0.62 | 0.53 | -0.02 | 0.01 | -1.40 | 0.16 |
| (4) | $\Delta PDS \leftrightarrow \Delta CT (\rho)$ | -0.03 | 0.01 | -2.68 | **<0.01** | -0.00 | 0.01 | -0.07 | 0.94 |
| **SA** |  |  |  |  |  |  |  |  |  |
| (1) | $PDS_{t1} \leftrightarrow SA_{t1} (\rho)$ | -0.00 | 0.01 | -0.04 | 0.97 | 0.04 | 0.01 | 3.64 | **<0.01** |
| (2) | $PDS_{t1} \rightarrow \Delta SA (\gamma_1)$ | -0.07 | 0.01 | -8.94 | **<0.01** | -0.03 | 0.01 | -3.60 | **<0.01** |
| (3) | $Area_{t1} \rightarrow \Delta PDS (\gamma_2)$ | 0.01 | 0.02 | 0.31 | 0.75 | -0.02 | 0.02 | -1.25 | 0.21 |
| (4) | $\Delta PDS \leftrightarrow \Delta SA (\rho)$ | 0.00 | 0.01 | 0.11 | 0.91 | 0.02 | 0.00 | 3.95 | **<0.01** |

**Note.** Table showing results from the multigroup BLCS model of Pubertal Development Scale (PDS) and MRI measures of Fractional Anisotropy (FA), Mean Diffusivity (MD), Cortical Thickness (CT), and Surface Area (SA), with estimates (Est), standard error (SE), z-score (z) and p-values (p). For each effect, $t_1$ represents baseline score, $\Delta PDS$ or $\Delta MRI$ represents the delta score in PDS and MRI measure between baseline and follow-up, ↔ represent correlations, and → represents regression coefficients. All reported effects are standardised. Bold fonts represent significant effects.

### 3.3. Pubertal development and fractional anisotropy

First, a BLCS for pubertal development and FA was modelled and the model fit was good: χ^2^ (28) = 272.45, CFI = 0.96, RMSEA = 0.04. Inspection of the four parameters of interest, reflecting the four possible puberty-brain relationships (outlined in Section 2.6), showed no statistically significant correlation between pubertal status and FA at baseline for males or females. The results did however show statistically significant associations of correlated change for both males (*r* = 0.03, *p* = 0.02) and females (*r* = 0.05, *p* < 0.01). In other words, those with greater increase in pubertal status were, on average, those with greater increase in FA. There were no coupling effects to report. Next, χ^2^-difference testing for sex differences were carried out. Here, no significant difference between males and females were found for any of the four puberty-brain relationships. For a graph representation of the results, see Figure 3.

### 3.4. Pubertal development and mean diffusivity

Second, a BLCS for pubertal development and MD was modelled and the model fit was good: χ^2^ (28) = 267.86, CFI = 0.96, RMSEA = 0.04. Inspection of the four parameters of interest revealed no statistically significant correlations or regression coefficients. χ^2^- difference tests revealed no significant difference between males and females for any of the four puberty-brain relationships. For a graph representation of the results, see Figure 4.

### 3.5. Pubertal development and cortical thickness

Third, a BLCS for pubertal development and cortical thickness was modelled and the model fit was good: χ^2^ (28) = 274.47, CFI = 0.96, RMSEA = 0.04. When examining the four parameters of interest, no statistically significant associations were observed between pubertal status and cortical thickness at baseline for males or females. The results did however reveal statistically significant correlated change for females (*r* = −0.03, *p* < 0.01), with no association present for males (*r* = −0.00, *p* = 0.94). In terms of coupling effects, pubertal status at baseline predicted rate of change in cortical thickness in females (*β* = −0.08, *p* < 0.01), while no associations were present in males. In other words, having initially higher PDS scores at baseline predicted larger increase between baseline and follow-up cortical thickness among females. χ^2^-difference tests revealed a significant difference between males and females ((df, 1) = [8.75], *p* < 0.01. *p^corr^*= 0.049) when comparing model parameters of pubertal status at baseline predicting rate of change in cortical thickness. The three other relationships investigated revealed no significant sex differences after correction for multiple comparisons. A graph representation of the results can be seen in Figure 5.

**Figure 5.**
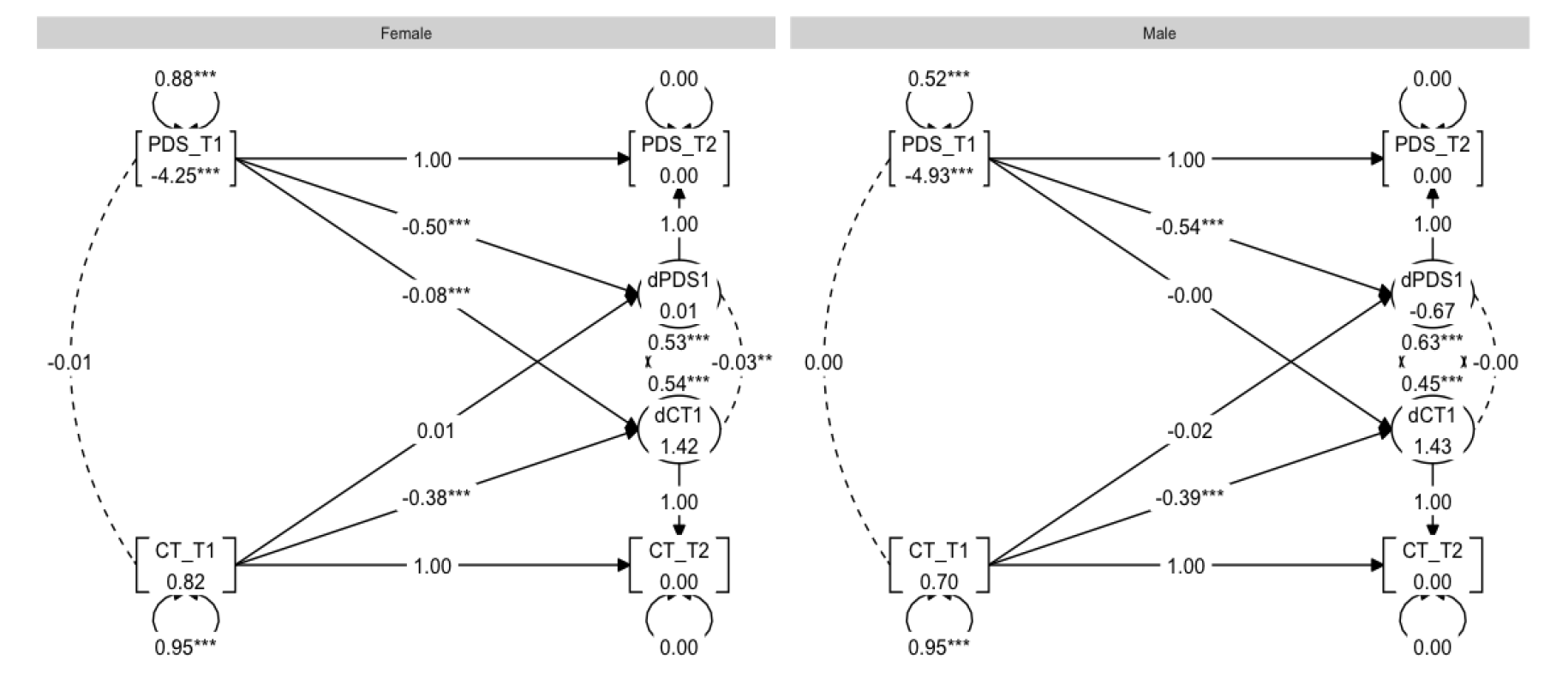
Graph showing the multigroup BLCS models for cortical thickness (CT) and PDS relations.

### 3.6. Pubertal development and surface area

Lastly, a BLCS for pubertal development and surface area was modelled and the model fit was good: χ^2^ (28) = 275.74, CFI = 0.98, RMSEA = 0.04. Inspection of the four parameters of interest showed statistically significant (positive) associations for the correlation between pubertal status and surface area at baseline (*r* = 0.04, *p* < 0.01) and correlated change for males (*r* = 0.02, *p* < 0.01), with no significant associations among females for either correlated baseline (*r* = −0.00, *p* = 0.97) or change (*r* = 0.00, *p* = 0.91). Coupling effects indicated pubertal status at baseline as being predictive of rate of change in surface area for both males (*β* = −0.03, *p* < 0.01) and females (*β* = −0.07, *p* < 0.01). In other words, having initially higher PDS scores at baseline predicted larger decrease between baseline and follow-up surface area measures. χ^2^-difference tests found a statistically significant sex difference ((df, 1) = [12.26], *p* < 0.01, *p^corr^* < 0.01) when comparing model parameters of pubertal status at baseline predicting rate of change in surface area, thereby showing that the association was significantly stronger for females than males. The three other relationships investigated revealed no significant group differences after correction for multiple comparisons. See Figure 6 for a graph representation of the results.

**Figure 6.**
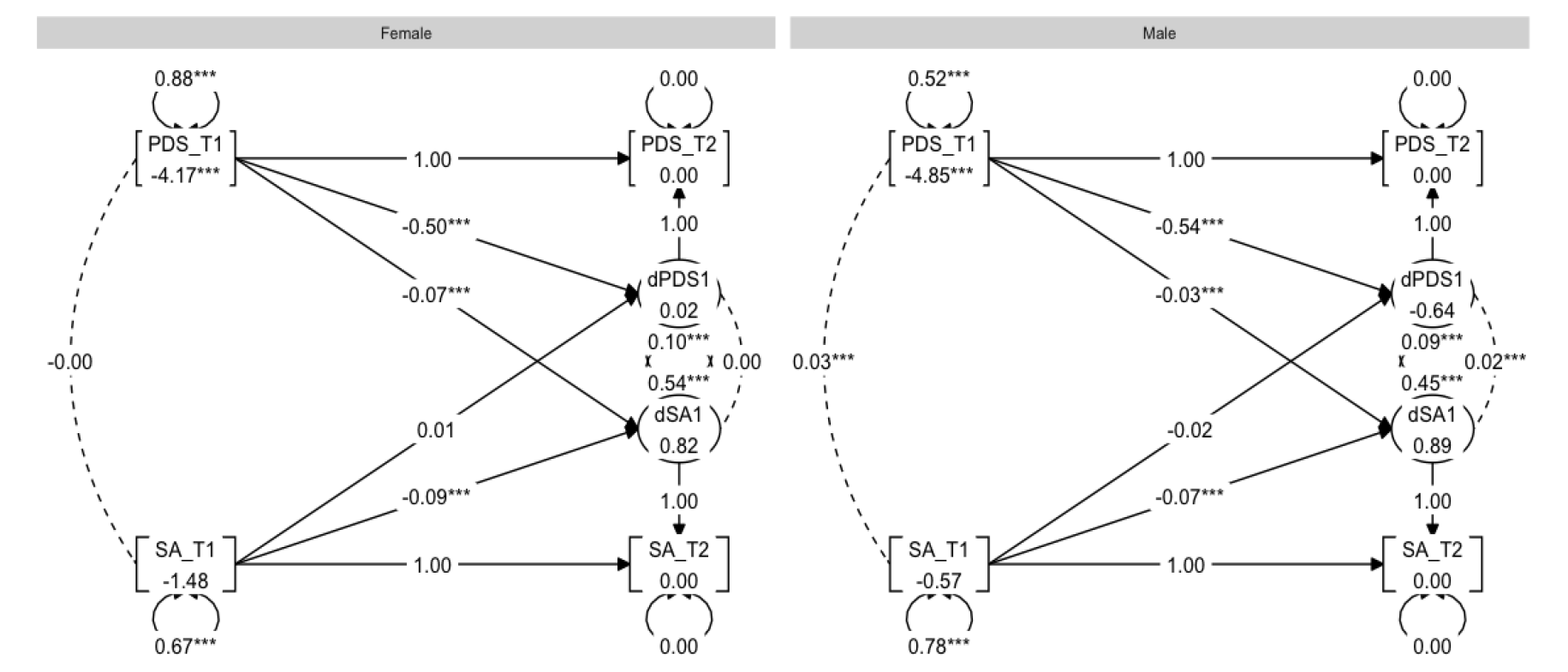
Graph showing the multigroup BLCS models for surface area (SA) and PDS relations.

## 4. Discussion

We investigated the coupling pathways of pubertal development and brain maturation in early adolescence using a multi-group BLCS framework in a large-scale longitudinal sample. Our main findings indicate that baseline pubertal status predicted the rate of change in cortical thickness for female youth only, and that pubertal status predicted rate of change in surface area for both sexes, but more strongly for females than males. Associations were also present for both correlation between pubertal status and surface area at baseline, and co-occurring changes over time, but these associations were present among male youth only. DTI analyses revealed correlated change over time between pubertal development and FA for both males and females, and no significant associations for MD. Overall, our results suggest sex-related differences in the associations between baseline pubertal status and rate of change in global cortical grey matter features of the brain.

Our study utilised a solid statistical framework to investigate the longitudinal coupling effects of pubertal development and structural brain maturation, providing valuable insight into the temporal dynamics of brain-and-puberty development. Previous research has reported sex-related associations between higher pubertal stage and global cortical grey matter (Herting et al., 2015), with studies generally reporting reduced cortical thickness in females (Bramen et al., 2011; Pfefferbaum et al., 2016) and no associations for males (Bramen et al., 2011; Koolschijn et al., 2014; Peper, Brouwer, et al., 2009). Our findings support previous findings and reveal additional knowledge into the coupling effects, with baseline pubertal status predicting rate of change in cortical thickness, as hypothesised.

For global surface area, baseline levels of pubertal status and surface area were positively related in males, going against our hypothesis. However, male participants who were more pubertally developed at baseline showed less increases in surface area compared to male participants who were less pubertally developed at baseline. Interestingly, our coupling effects provide evidence suggesting that pubertal status at baseline predicts rate of change in surface area (for both males and females), with faster pubertal tempo associated with greater decrease in surface area, supporting our hypothesis. Moreover, despite both sexes showing effects, our findings revealed significantly stronger effects in females than males.

For surface area, our findings can be best understood in the context of the age range in this sample. While the directionality of the coupling effects go in the expected directions (i.e., more pubertally developed predicting surface area *decrease*), one unexpected finding is the observation that males show the opposite trend for baseline levels of pubertal status and surface area (i.e., positively related). Previous research has reported decreases in surface area associated with more pubertally developed males (Herting et al., 2015). One possible explanation for this could be that many of the males in the present sample are still experiencing increase in surface area (see e.g., Raznahan et al. (2011) and SI Figure 7).

Additionally, as evidenced by Figure 1 and PDS descriptive statistics in Table 1, the majority of the males in our sample are in the early stages of pubertal development compared to females (mid-late stages), and do not progress a lot throughout the study period, with the average PDS moving from 7.5 at baseline to 8.8 at follow-up. This is a major limitation of the current study. ABCD Study data collection is ongoing and thus, the age range available at the time of analysis is not representative of the entire pubertal period. While the current study is an important first step in investigating the emergence of sex differences in pubertal and brain development, longitudinal data spanning the entire range of puberty will be needed to advance our understanding of sex-specific developmental patterns.

Previous research investigating associations between pubertal development and DTI is sparse. The one existing longitudinal DTI study (Herting et al. 2017) reported that increase in PDS was associated with increase in FA, which was supported in our study using a much larger sample. However, while the prior study reported sex-related effects, our results did not indicate differences between males and females. One potential explanation for this is that Herting and colleagues looked at various regions of the brain while our study observed global FA, meaning if a minority of regional effects (as observed in Herting et al. (2017)) were going the other direction, they may not be evident in the overall effect of increasing FA in the majority of other regions. No evidence of correlation at baseline or coupling effects were observed in either FA or MD, and there were no significant sex differences, which goes against our previously stated hypotheses.

Adolescence is a sensitive period that offers a window of opportunity to shape the fate of the adult brain. Within that period, accelerated pubertal development has been identified as a potential indicator of concern for the developing brain, with earlier pubertal timing being linked to psychological outcomes such as depression, anxiety, and ADHD (Copeland et al., 2019; Ge et al., 2006; Mendle et al., 2010; Mendle & Ferrero, 2012). Future research should focus on increasing our understanding on the extent to intervention strategies targeting early pubertal-timing-related risk factors reduce the risk of related psychopathology. Explicitly investigating the brain-puberty relationship with sex differences in mind is also important for future research when considering that risk of worse mental health outcomes for females and males may differ, and this may have ramifications for related brain changes.

With research showing earlier pubertal timing in those facing barriers to positive health and well-being, questions must be raised of how to tackle the challenges posed by environmental factors and disparities in living conditions. Research has observed earlier pubertal timing in concordance with higher child weight status, lower SES, and nutritional needs not being met, and these have disproportionately impacted minority groups more. Although speculative, environmental pressures may be central agents in advanced adaptation to adult roles in the environment, termed ‘adultification’ (Pfeifer & Allen, 2021), where adolescents may be undergoing earlier onset and tempo of puberty, and related neurodevelopmental changes, as a means to adapt to a harsh environment, subsequently increasing their risk of psychopathology. More research is needed to elucidate the potential role of environmental factors on advanced pubertal development and brain maturation.

The current study has several strengths and limitations that warrant further discussion. Firstly, the ABCD Study cohort provides a well-powered sample that is openly available for researchers, improving both transparency and encouraging initiatives testing the robustness of the findings. Second, the longitudinal data and analyses allow for tracking changes over time, albeit while still limited by only one follow-up assessment. Nevertheless, the current multigroup BLCS analysis framework is well tailored to address hypotheses related to both general and fine-grained temporal dynamics over time and is especially powerful for testing cross-domain couplings (Kievit et al., 2018).

While global measures may capture broad effects, we know that pubertal development is associated with regional changes in frontal, temporal and parietal areas (as well as subcortical regions) which provides rationale for looking at these in future research. FA and MD revealed fewer effects compared to grey matter morphology. Future research should utilise advanced diffusion MRI metrics (Beck et al., 2021; Tamnes et al., 2018) that differentiate between intra-and extra-axonal compartments and potentially provides more information than conventional DTI. Lastly, future research would benefit from a combined approach of assessing pubertal development by means of questionnaires *as well as* hormonal data, given the limitations of PDS (Cheng et al., 2021).

Some general limitations pertaining to ABCD Study and other large-scale volunteer scale databases include biases related to being more healthy, more wealthy, and more educated compared to the general population in their respective countries (Brayne & Moffitt, 2022). This lack of representativeness may mean that those with most adversity and disadvantage will not be represented in this research. Future research should consider more diversified samples to reduce the healthy volunteer bias compromising the generalisability of the findings to more vulnerable populations. On the other hand, the ABCD Study cohort does include more ethnically diverse participants than similar large-scale volunteer databases (Brayne & Moffitt, 2022). Due to participant ages at recruitment, the ABCD Study data does not capture early gonadal processes for many participants (Cheng et al., 2021; Herting et al., 2021), which is more pronounced for females and non-White participants. We also acknowledge that the ABCD Study cohort shows some overlap in age and PDS between the included timepoints in the study (see Figure 1), and thus caution must be taken in interpreting results. Additionally, we recognise that variations in pubertal development in, for example, early stages vs mid-late stages may differ in their associations to brain maturation, and such possible non-linear associations were not examined in the current study. Future research would benefit from analysing samples with multiple timepoints and an age-range spanning the entire span of pubertal development and utilising longitudinal modelling that accommodates potentially non-linear relationships between brain maturation and pubertal development.

In conclusion, our analyses highlight pubertal status in early adolescence as being predictive of longitudinal changes in cortical grey matter structure over time, and that these changes are different for males and females. Further research is needed to understand how puberty impacts the brain, the contribution of environmental and lifestyle factors on pubertal development and brain maturation, and, in turn, the effects on mental health.

## Data Availability

Data used in the present study were drawn from the ABCD curated annual release 4.0, containing data from baseline up until the second-year visit (https://data-archive.nimh.nih.gov/abcd). All ABCD Study data is stored in the NIMH Data Archive Collection #2573, which is available for registered and authorised users.

## Acknowledgments

Data used in the preparation of this article were obtained from the Adolescent Brain Cognitive DevelopmentSM (ABCD) Study (https://abcdstudy.org), held in the NIMH Data Archive (NDA). This is a multisite, longitudinal study designed to recruit more than 10,000 children aged 9-10 and follow them over 10 years into early adulthood. The ABCD Study® is supported by the National Institutes of Health and additional federal partners under award numbers U01DA041048, U01DA050989, U01DA051016, U01DA041022, U01DA051018, U01DA051037, U01DA050987, U01DA041174, U01DA041106, U01DA041117, U01DA041028, U01DA041134, U01DA050988, U01DA051039, U01DA041156, U01DA041025, U01DA041120, U01DA051038, U01DA041148, U01DA041093, U01DA041089, U24DA041123, U24DA041147. A full list of supporters is available at https://abcdstudy.org/federal-partners.html. A listing of participating sites and a complete listing of the study investigators can be found at https://abcdstudy.org/consortium_members/. ABCD consortium investigators designed and implemented the study and/or provided data but did not necessarily participate in the analysis or writing of this report. This manuscript reflects the views of the authors and may not reflect the opinions or views of the NIH or ABCD consortium investigators.

