## Supplementary Material for "Puberty differentially predicts brain maturation in male and female youth: A longitudinal ABCD Study"

**SI Section 1.** Quality control

For MRI data, quality check procedures followed the standard protocol described in Hagler et al. (2019). Briefly, participants with excessive head motion or poor data quality were excluded from the curated data release by the ABCD team. Additional quality assurance was carried out following extraction of data using the recommendations for data cleaning provided by the ABCD team (using data structure abcd_imgincl01). Here, recommendation criteria for include/exclude is marked by 1/0, respectively, and removal of data marked 0 was carried out. Following this procedure, cortical gray matter data was reduced from 19,586 observations (obs) to 19,417 and DTI data was reduced from 19,587 obs to 18,847. Next, during exclusion of siblings to minimize confounding effects from family-related factors, the datasets reduced to 17,492 obs for cortical gray matter and 17,046 obs for DTI. Next, brain measures were inspected manually for any additional outliers by checking for values ± 5 SD and using histogram figures for visualisation (SI Figures 2-3 below). During this manual inspection data cleaning, the cortical data remained at 17,492 obs and the DTI data was reduced to 16,620 obs. The cortical and DTI brain features of interest were then merged together, followed by being merged with quality checked total PDS and covariates of interest (Section 2.7 and SI Section 3). Due to LongCombat’s (Section 2.5) inability to deal with missing data, all remaining NAs were also removed during this step, reducing the dataset to 14,995 obs including our final sample of 8,896 participants at baseline and 6,099 participants at follow up.

An attrition analysis was carried out on the number of obs in the dataset, comparing N obs of the initial sample (19,587) and N obs for the final sample (14,995). The results of the analysis revealed an attrition rate of 23.44% (*minus 4592 obs*). An attrition analysis of the sample (participants) was also carried out, using the unique IDs of the initial (N = 11,811) and final (N = 8,896) datasets. The analysis revealed an attrition rate of 13.53% *(minus 2,915 participants)*. Finally, an attrition analysis was carried out to test the rate of drop out of participants between baseline and follow-up, including testing for differences in characteristics of baseline participants that were retained for follow-up versus those that dropped out. The analysis revealed an attrition rate of 31.4% (2797 participants that were not retained between baseline and follow-up from the final sample of 8,896). SI Figure 1 shows characteristics of retained versus non-retained participants, including differences in age, BMI, SES, and PDS.

For PDS, raw data was examined and participants with missing data or ‘I do not know’ (coded 999) were removed from the data frame. The remaining participants were valid for inclusion if they had data for each of the five items of the PDS (see Section 2.6). Following data cleaning, a total PDS score was calculated, which was subsequently used for the analysis.

For covariates of genetic ancestry-derived ethnicity, BMI, and SES, data cleaning involved removal of participants with missing data and manual inspection of outliers. BMI values over 50 or less than 10 were determined extreme cases based on BMI-for-age growth charts and recommendations by Centres for Disease Control and Prevention (CDC) (US Preventive Services Task Force, 2017). For SES and genetic ancestry-derived ethnicity, no additional data (outliers) were removed following removal of missing data. For visualisation of distribution pertaining to BMI and SES, see SI Figures 4-5.

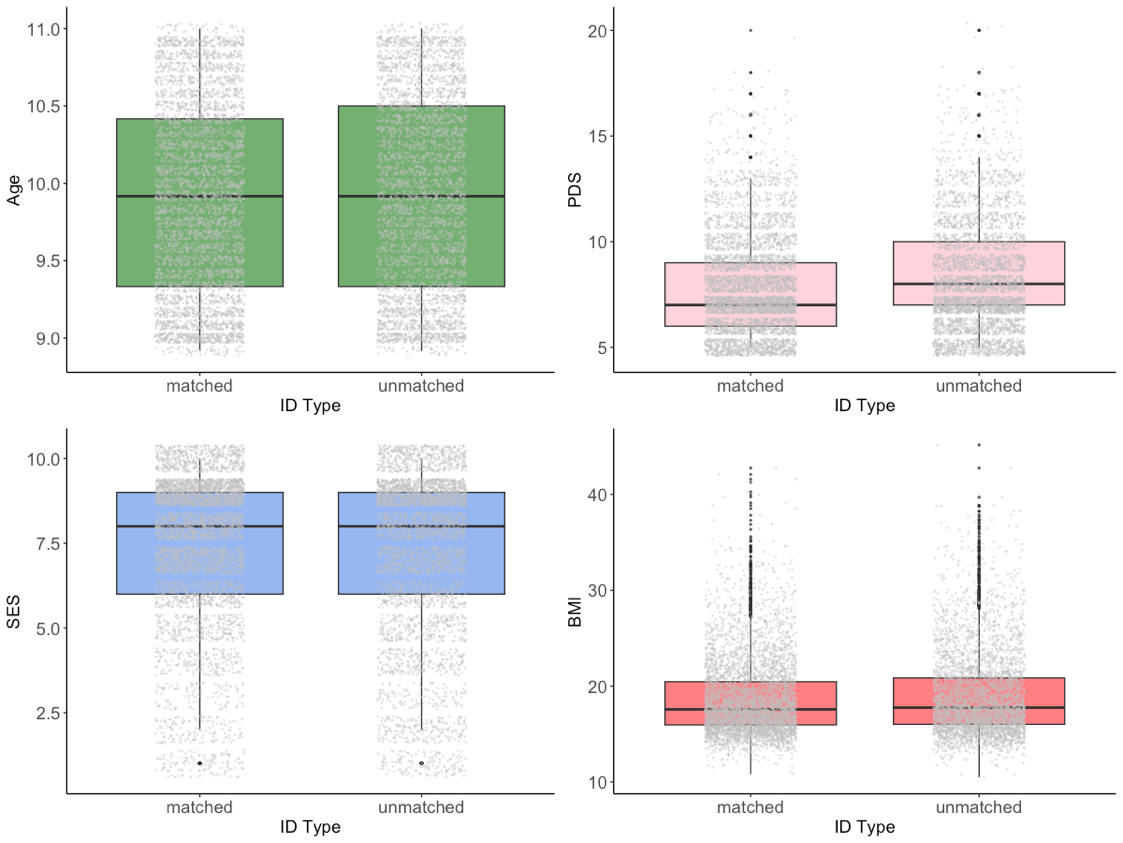

**SI Figure 1**. Boxplots showing retained (matched) and non-retained (unmatched) participants. Each boxplot compares baseline measures of age, PDS, SES, and BMI scores for each group.

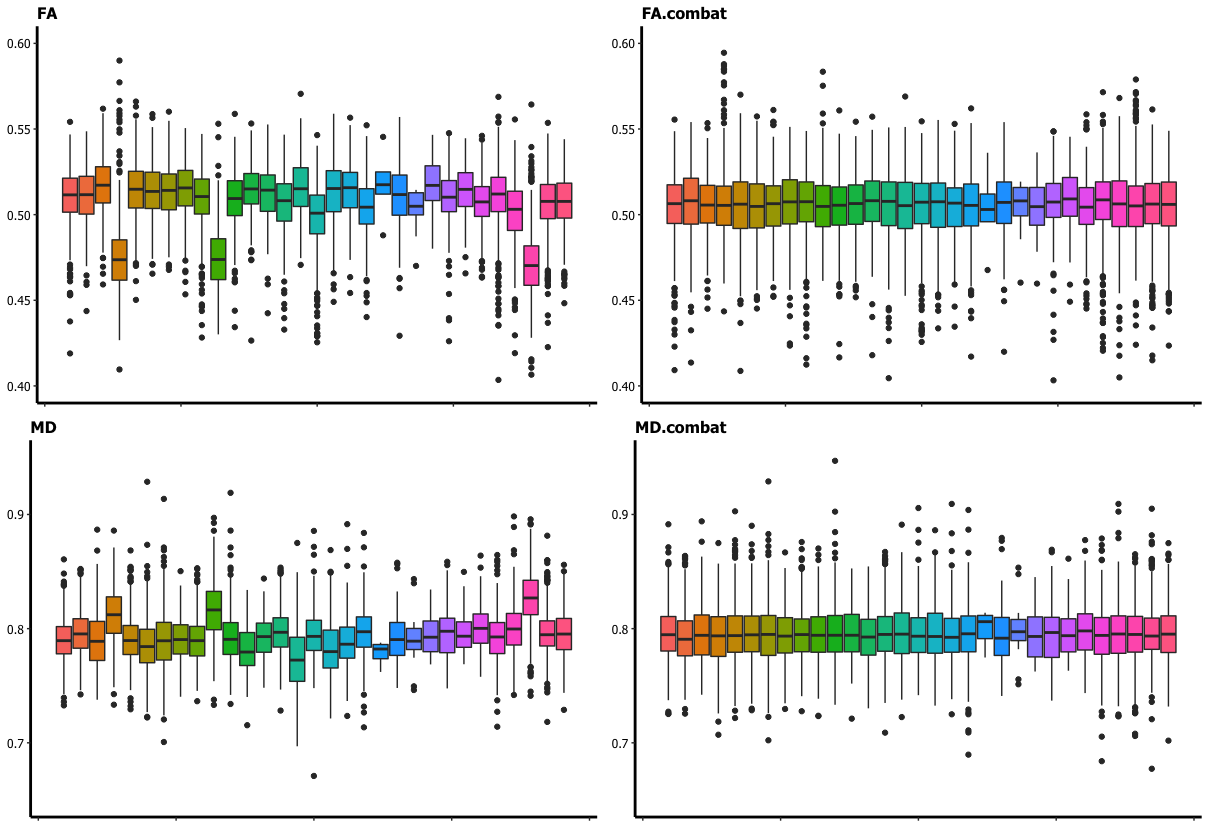

**SI Figure 2**. Distribution of FA and MD residuals across scanners before harmonisation (left), and after longitudinal ComBat (right). For each model, age, BMI, SES, and genetic ancestry-derived ethnicity are added as covariates. Each colour represents a different scanner within each figure, with the scanner from non-harmonised data matching combatted data in colour.

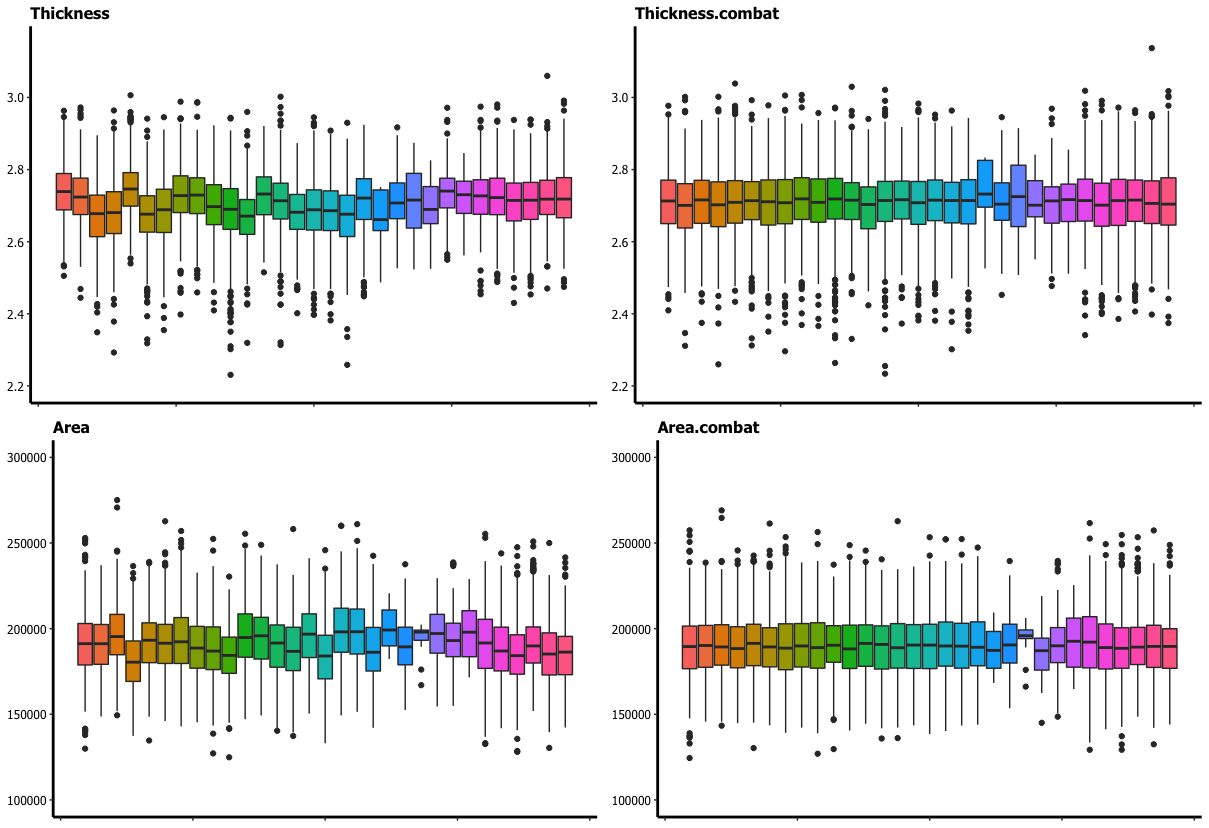

**SI Figure 3**. Distribution of cortical thickness and surface area residuals across scanners before harmonisation (left), and after longitudinal ComBat (right). For each model, age, BMI, SES, and genetic ancestry-derived ethnicity are added as covariates. Each colour represents a different scanner within each figure, with the scanner from non-harmonised data matching combatted data in colour.

**SI Section 2.** Ethical approval

The Institutional Review Board (IRB) at the University of California, San Diego, approved all aspects of the ABCD Study (Auchter et al., 2018). Parents or guardians provided written consent, while the child provided written assent. The current study was conducted in line with the Declaration of Helsinki and was approved by the Regional Committee for Medical and Health Research Ethics (REK 2019/943).

**SI Section 3.** MRI acquisition and processing

Cortical reconstruction and volumetric segmentation was performed with FreeSurfer 6.0 (Dale et al., 1999; Fischl et al., 2002). This processing includes motion correction and averaging (Reuter et al., 2010), removal of non-brain tissue (Ségonne et al., 2004), automated Talairach transformation, segmentation of the subcortical white matter and deep gray matter volumetric structures (Fischl et al., 2002, 2004), intensity normalization (Sled et al., 1998), tessellation of the gray matter white matter boundary, automated topology correction (Fischl et al., 2001; Ségonne et al., 2007), and surface deformation (Dale et al., 1999).

White matter microstructural measures were generated using AtlasTrack, a probabilistic atlas-based method for automated segmentation of white matter fiber tracts (Hagler Jr. et al., 2009), as described in (Hagler et al., 2019). Briefly, structural MRI images for each subject are nonlinearly registered to the atlas using discrete cosine transforms (Friston et al., 1995), and DTI-derived diffusion orientations for each subject are compared to the atlas fiber orientations. Voxels containing primarily gray matter or cerebral spinal fluid, identified using FreeSurfer’s automated brain segmentation (Fischl et al., 2002), are excluded from analysis.

**SI Section 4.** Covariates of interest

BMI was derived by dividing an individual’s weight in kilograms (kg) by their height in metres squared (m^2^). SES was calculated using the average score from three related income measures; parental income, partner income, and combined income, resulting in a score that represent the total household income. For SES, categories between 1-7 represented the following income bracket: 1 = Less than $5,000; 2 = $5,000 through $11,999; 3 = $12,000 through $15,999; 4 = $16,000 through $24,999; 5 = $25,000 through $34,999; 6 = $35,000 through $49,999; 7 = $50,000 through $74,999; 8 = $75,000 through $99,999; 9 = $100,000 through $199,999; 10 = $200,000 and greater. For genetic ancestry-derived ethnicity, four continuous variables (0-1) labelled African, American, European, and East Asian were utilised, which are based on calculations of ethnicity using Bayesian clustering from genetic data with 1000 genome reference panel (Raj et al., 2014). Figures pertaining to each covariate and the associations between them can be found in SI Figures 4, 5, and 6.

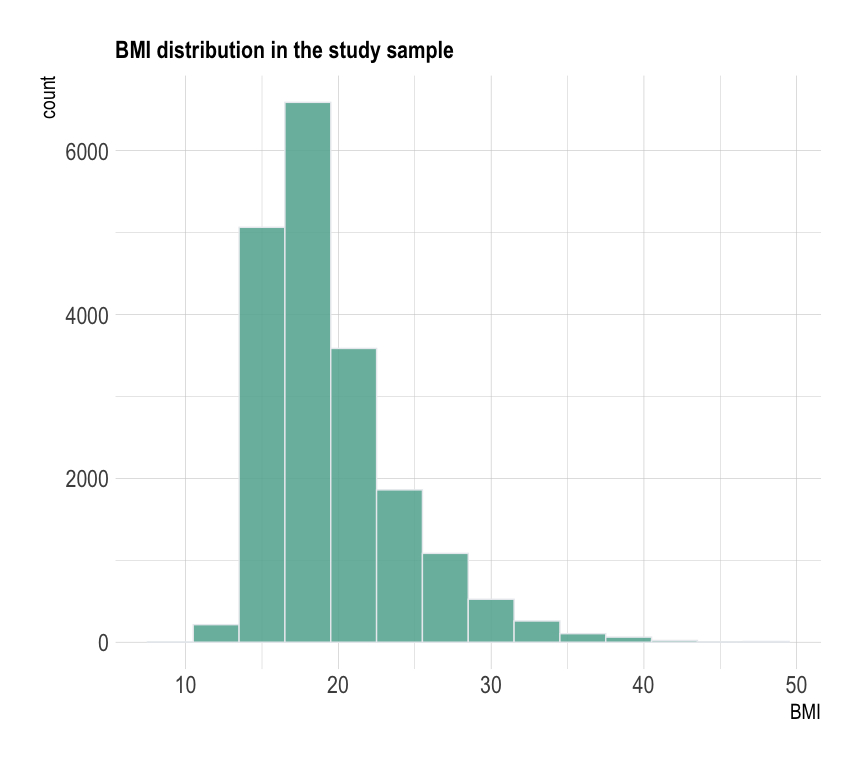

**SI Figure 4.** Histogram figure showing distribution of BMI scores for the full study sample, calculated by dividing an individual’s weight in kilograms (kg) by their height in metres squared (m^2^).

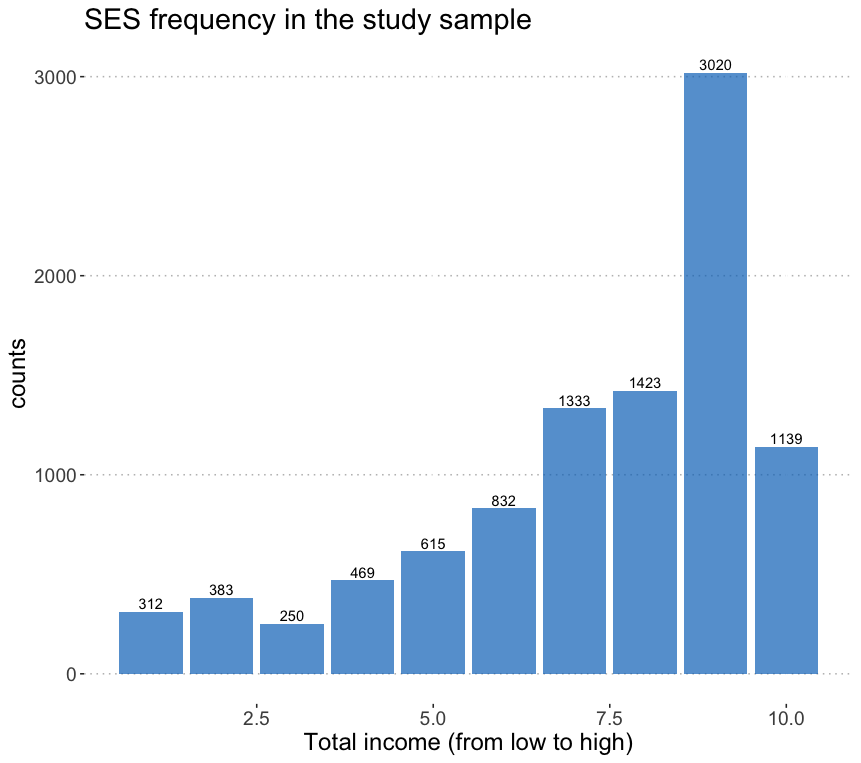

**SI Figure 5.** Figure showing frequency distribution of SES scores for the full study sample, including count (N participants) belonging to each SES category. 1 = Less than $5,000; 2 = $5,000 through $11,999; 3 = $12,000 through $15,999; 4 = $16,000 through $24,999; 5 = $25,000 through $34,999; 6 = $35,000 through $49,999; 7 = $50,000 through $74,999; 8 = $75,000 through $99,999; 9 = $100,000 through $199,999; 10 = $200,000 and greater.

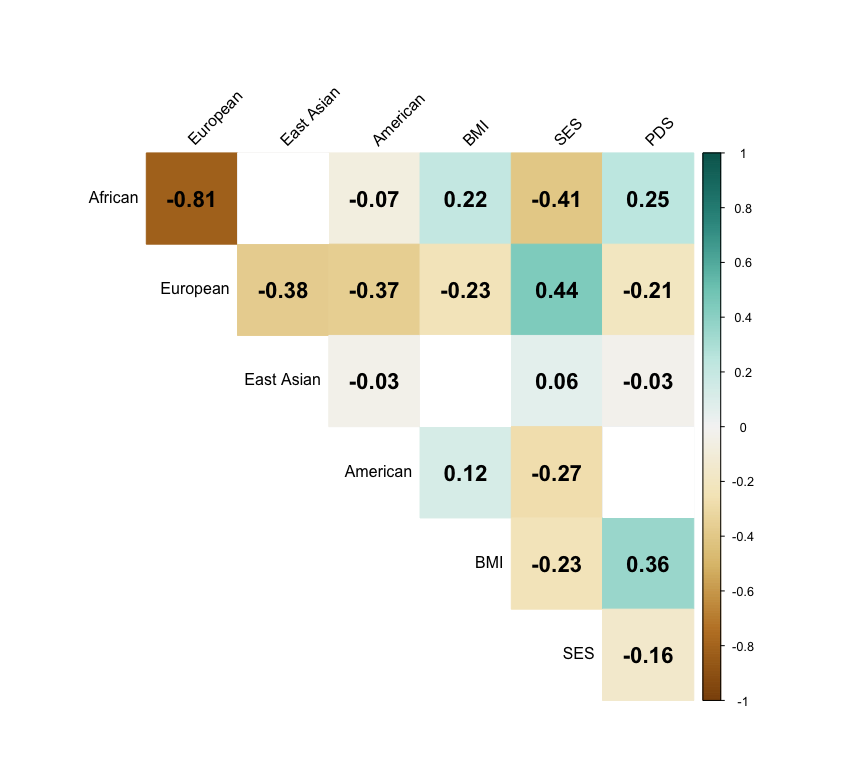

**SI Figure 6.** Correlation matrix showing relatedness between covariates of interest included in the study analysis. Ethnicity data is described proportionally from genetic data derived as continuous variables between zero and one and labelled African, American, European, and East Asian.

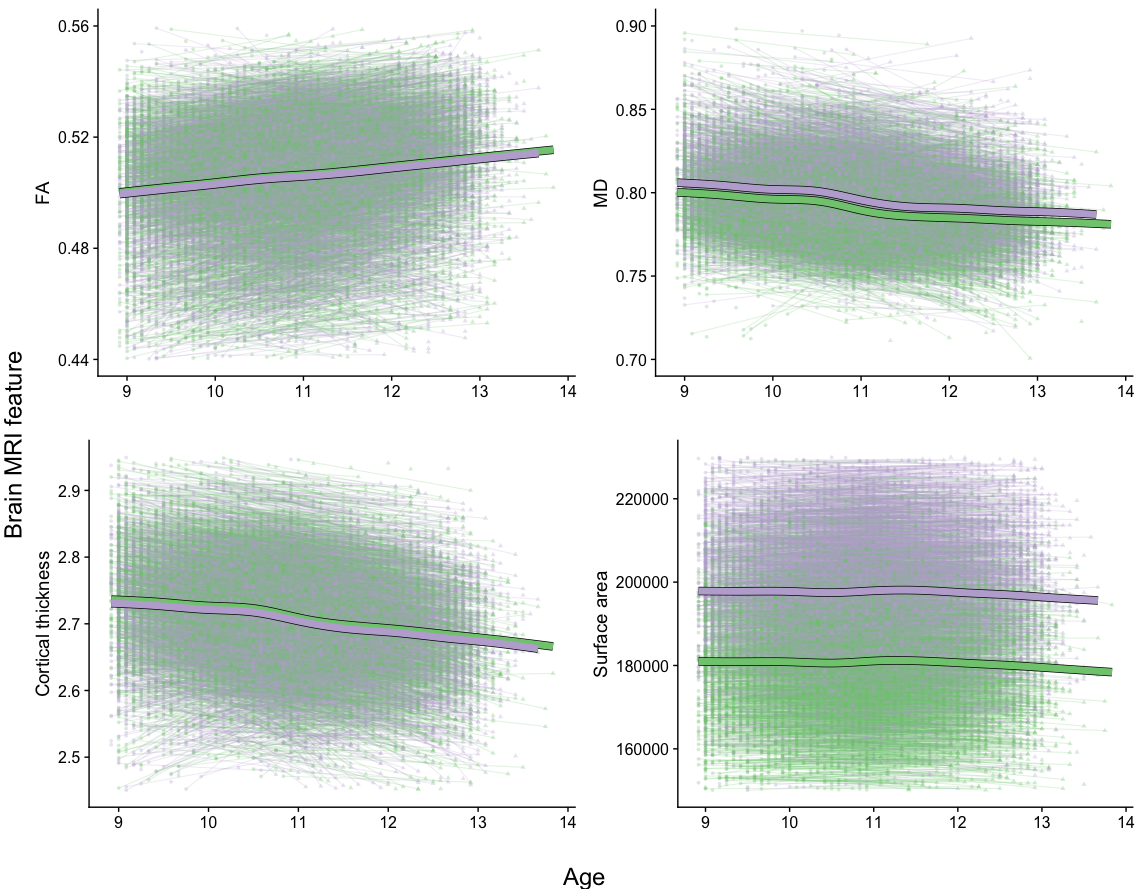
 **SI Figure 7.** Age curves where each brain MRI feature (y-axis) is plotted as a function of age (x-axis). Fitted lines are made with linear mixed effects (lme) model-derived predicted values. Points connected by lines represent longitudinal data where circles are baseline and triangles are follow-up assessments. Male subjects are represented by lilac and female subjects by green.

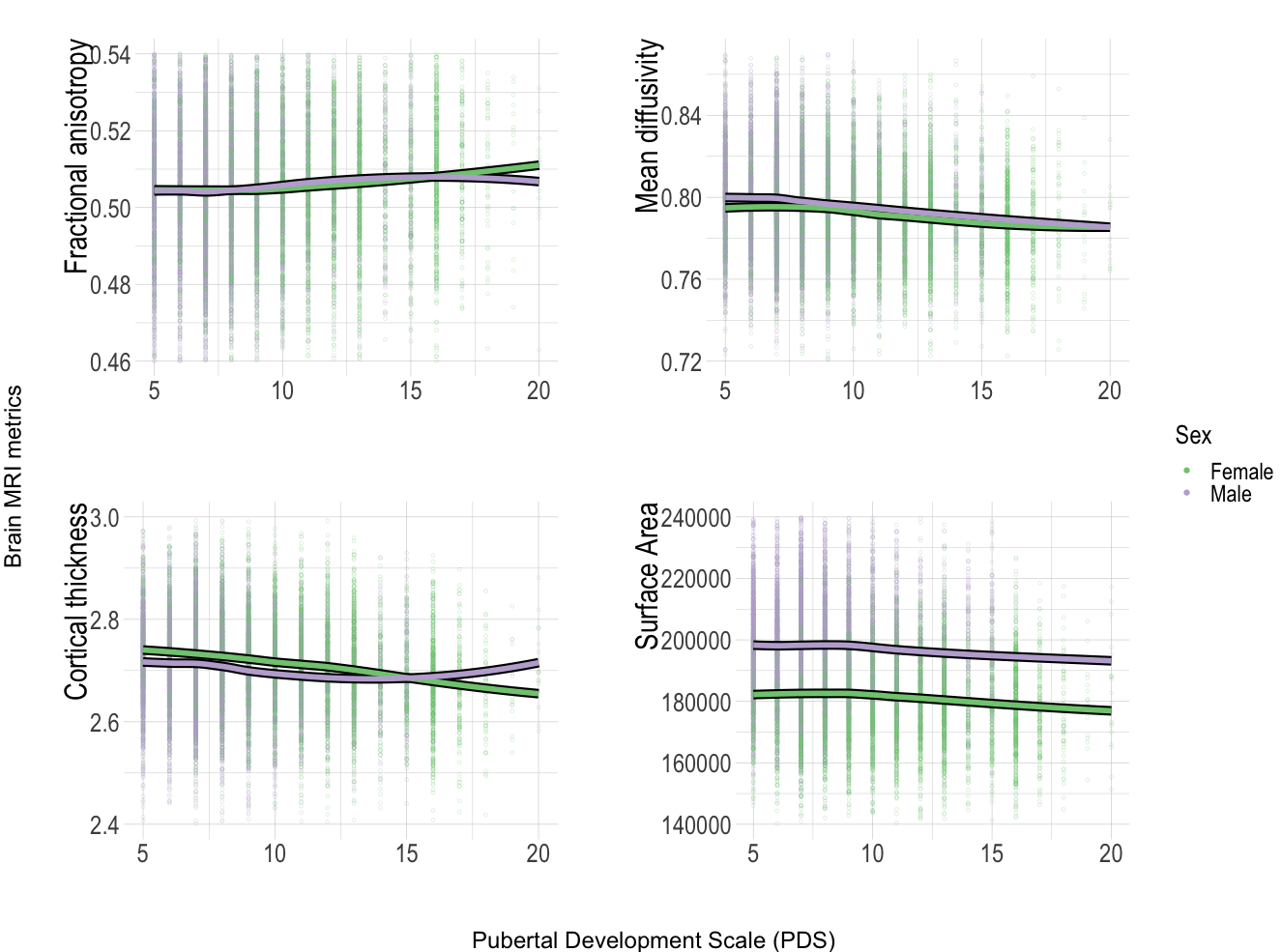

**SI Figure 8**. Brain MRI metrics as a function of Pubertal Development Scale (PDS) score.

**SI Section 5.** Multigroup bivariate latent change score (BLCS) model. This section outputs the full results of our BLCS model created in R markdown with notes added for clarification of model.

### **Multi-group Bivariate Latent Change Score (BLCS) models**

#### **Analysis overview**

In this document, we look at some of the output from four different multi-group BLCS models - one for each brain MRI modality. All models are updated with covariates of age (with fixed parameters indicated by added vectors multiplied by e.g., age: c(a,a)*age). This ensures that the two parameters “e.g. (a,a)” are now constrained to be equal across the two groups (male, female). Additionally, body-mass index (BMI), Genetic Ancestry factor for African, East Asian, European, American, socioeconomic status (SES) are also added as covariates and constrained to be equal across the two groups.

###### **Covariates**

Covariates have been added using the following lines of code in each model:

- dPDS1 ~ c(a,a)* age + c(a2,a2)* BMI + c(a3,a3)* SES + c(a4,a4)* gen_african + c(a5,a5)* gen_european + c(a6,a6)* gen_east_asian + c(a7,a7)* gen_american
- PDS_T1 ~ c(b,b)* age + c(b2,b2)* BMI + c(b3,b3)* SES + c(b4,b4)* gen_african + c(b5,b5)* gen_european + c(b6,b6)* gen_east_asian + c(b7,b7)* gen_american
- dFA1 ~ c(c,c)* age + c(c2,c2)* BMI + c(c3,c3)* SES + c(c4,c4)* gen_african + c(c5,c5)* gen_european + c(c6,c6)* gen_east_asian + c(c7,c7)* gen_american
- FA_T1 ~ c(d,d)* age + c(d2,d2)* BMI + c(d3,d3)* SES + c(d4,d4)* gen_african + c(d5,d5)* gen_european + c(d6,d6)* gen_east_asian + c(d7,d7)* gen_american

**BLCS Fractional Anisotropy**

First, we run the model investigating white matter microstructural FA and pubertal maturation.

| **Model Significance** | | | |
| --- | --- | --- | --- |
| **Sample.Size** | **Chi.Square** | **df** | **p.value** |
| 9195 | 272.449 | 28 | 0 |

| **Model Fit Measures** | | | | | |
| --- | --- | --- | --- | --- | --- |
| **CFI** | **RMSEA** | **RMSEA.Lower** | **RMSEA.Upper** | **AIC** | **BIC** |
| 0.956 | 0.044 | 0.039 | 0.048 | 66202.68 | 66601.76 |

| **Covariances** | | | | | | | | | | |
| --- | --- | --- | --- | --- | --- | --- | --- | --- | --- | --- |
| lhs | op | rhs | group | label | est.std | se | z | pvalue | ci.lower | ci.upper |
| PDS_T1 | ~~ | FA_T1 | 1 |  | -0.00 | 0.02 | -0.27 | 0.79 | -.038 | .029 |
| dPDS1 | ~~ | dFA1 | 1 |  | 0.05 | 0.01 | 3.54 | 0.00 | .036 | .122 |
| PDS_T1 | ~~ | FA_T1 | 2 |  | 0.00 | 0.01 | 0.14 | 0.89 | -.029 | .034 |
| dPDS1 | ~~ | dFA1 | 2 |  | 0.03 | 0.01 | 2.36 | 0.02 | .008 | .088 |

| **Factor Loadings** | | | | | | | | |
| --- | --- | --- | --- | --- | --- | --- | --- | --- |
|  |  | **Standardized** | | | | | | |
| **Latent Factor** | **Indicator** | **Loadings** | **sig** | **p** | **Lower.CI** | **Upper.CI** | **SE** | **z** |
| dPDS1 | PDS_T2 | 0.941 | *** | 0 | 0.904 | 0.979 | 0.019 | 49.201 |
| dFA1 | FA_T2 | 1.006 | *** | 0 | 0.947 | 1.065 | 0.030 | 33.570 |
| dPDS1 | PDS_T2 | 0.979 | *** | 0 | 0.938 | 1.021 | 0.021 | 46.171 |
| dFA1 | FA_T2 | 0.951 | *** | 0 | 0.893 | 1.008 | 0.029 | 32.453 |

| **Regression Paths** | | | | | | | | |
| --- | --- | --- | --- | --- | --- | --- | --- | --- |
|  |  | **Standardized** | | | | | | |
| **Predictor** | **DV** | **Path Values** | **SE** | **z** | **sig** | **p** | **Lower.CI** | **Upper.CI** |
| PDS_T1 | PDS_T2 | 1.107 | 0.015 | 72.996 | *** | 0.000 | 1.077 | 1.136 |
| FA_T1 | FA_T2 | 1.039 | 0.027 | 38.177 | *** | 0.000 | 0.985 | 1.092 |
| FA_T1 | dPDS1 | -0.016 | 0.019 | -0.831 |  | 0.406 | -0.054 | 0.022 |
| PDS_T1 | dPDS1 | -0.587 | 0.014 | -42.073 | *** | 0.000 | -0.614 | -0.560 |
| PDS_T1 | dFA1 | -0.001 | 0.020 | -0.066 |  | 0.947 | -0.041 | 0.039 |
| FA_T1 | dFA1 | -0.527 | 0.023 | -22.620 | *** | 0.000 | -0.572 | -0.481 |
| age | dPDS1 | 0.074 | 0.012 | 6.299 | *** | 0.000 | 0.051 | 0.097 |
| BMI | dPDS1 | 0.102 | 0.013 | 7.876 | *** | 0.000 | 0.076 | 0.127 |
| SES | dPDS1 | 0.002 | 0.014 | 0.155 |  | 0.877 | -0.025 | 0.029 |
| gen_african | dPDS1 | -0.230 | 0.322 | -0.714 |  | 0.475 | -0.861 | 0.401 |
| gen_european | dPDS1 | -0.291 | 0.355 | -0.820 |  | 0.412 | -0.986 | 0.404 |
| gen_east_asian | dPDS1 | -0.074 | 0.105 | -0.703 |  | 0.482 | -0.279 | 0.132 |
| gen_american | dPDS1 | -0.093 | 0.143 | -0.649 |  | 0.516 | -0.373 | 0.187 |
| age | PDS_T1 | 0.138 | 0.013 | 10.549 | *** | 0.000 | 0.112 | 0.164 |
| BMI | PDS_T1 | 0.196 | 0.011 | 17.821 | *** | 0.000 | 0.175 | 0.218 |
| SES | PDS_T1 | -0.095 | 0.012 | -8.262 | *** | 0.000 | -0.118 | -0.073 |
| gen_african | PDS_T1 | 0.874 | 0.262 | 3.330 | *** | 0.001 | 0.359 | 1.388 |
| gen_european | PDS_T1 | 0.718 | 0.290 | 2.474 |  | 0.013 | 0.149 | 1.287 |
| gen_east_asian | PDS_T1 | 0.193 | 0.086 | 2.254 |  | 0.024 | 0.025 | 0.360 |
| gen_american | PDS_T1 | 0.293 | 0.116 | 2.524 |  | 0.012 | 0.065 | 0.520 |
| age | dFA1 | 0.003 | 0.014 | 0.212 |  | 0.832 | -0.024 | 0.030 |
| BMI | dFA1 | -0.022 | 0.016 | -1.389 |  | 0.165 | -0.054 | 0.009 |
| SES | dFA1 | 0.044 | 0.016 | 2.699 | ** | 0.007 | 0.012 | 0.077 |
| gen_african | dFA1 | -0.368 | 0.346 | -1.063 |  | 0.288 | -1.046 | 0.310 |
| gen_european | dFA1 | -0.380 | 0.380 | -0.998 |  | 0.318 | -1.125 | 0.366 |
| gen_east_asian | dFA1 | -0.113 | 0.112 | -1.012 |  | 0.311 | -0.333 | 0.106 |
| gen_american | dFA1 | -0.172 | 0.153 | -1.121 |  | 0.262 | -0.473 | 0.129 |
| age | FA_T1 | 0.130 | 0.017 | 7.627 | *** | 0.000 | 0.096 | 0.163 |
| BMI | FA_T1 | 0.004 | 0.013 | 0.316 |  | 0.752 | -0.022 | 0.030 |
| SES | FA_T1 | 0.014 | 0.014 | 0.946 |  | 0.344 | -0.015 | 0.042 |
| gen_african | FA_T1 | -0.379 | 0.336 | -1.127 |  | 0.260 | -1.037 | 0.280 |
| gen_european | FA_T1 | -0.331 | 0.370 | -0.894 |  | 0.371 | -1.057 | 0.395 |
| gen_east_asian | FA_T1 | -0.110 | 0.109 | -1.011 |  | 0.312 | -0.323 | 0.103 |
| gen_american | FA_T1 | -0.126 | 0.149 | -0.850 |  | 0.395 | -0.418 | 0.165 |
| PDS_T1 | PDS_T2 | 1.028 | 0.019 | 53.370 | *** | 0.000 | 0.990 | 1.066 |
| FA_T1 | FA_T2 | 1.003 | 0.022 | 46.327 | *** | 0.000 | 0.960 | 1.045 |
| FA_T1 | dPDS1 | 0.009 | 0.018 | 0.497 |  | 0.619 | -0.026 | 0.044 |
| PDS_T1 | dPDS1 | -0.568 | 0.018 | -31.452 | *** | 0.000 | -0.604 | -0.533 |
| PDS_T1 | dFA1 | 0.000 | 0.021 | -0.004 |  | 0.997 | -0.041 | 0.041 |
| FA_T1 | dFA1 | -0.481 | 0.023 | -20.810 | *** | 0.000 | -0.526 | -0.435 |
| age | dPDS1 | 0.084 | 0.013 | 6.297 | *** | 0.000 | 0.058 | 0.110 |
| BMI | dPDS1 | 0.106 | 0.013 | 7.901 | *** | 0.000 | 0.080 | 0.133 |
| SES | dPDS1 | 0.002 | 0.015 | 0.155 |  | 0.877 | -0.028 | 0.033 |
| gen_african | dPDS1 | -0.244 | 0.341 | -0.715 |  | 0.475 | -0.913 | 0.425 |
| gen_european | dPDS1 | -0.330 | 0.402 | -0.820 |  | 0.412 | -1.118 | 0.458 |
| gen_east_asian | dPDS1 | -0.126 | 0.180 | -0.703 |  | 0.482 | -0.478 | 0.226 |
| gen_american | dPDS1 | -0.117 | 0.180 | -0.650 |  | 0.516 | -0.470 | 0.236 |
| age | PDS_T1 | 0.176 | 0.016 | 10.785 | *** | 0.000 | 0.144 | 0.208 |
| BMI | PDS_T1 | 0.230 | 0.012 | 18.412 | *** | 0.000 | 0.206 | 0.255 |
| SES | PDS_T1 | -0.120 | 0.014 | -8.433 | *** | 0.000 | -0.148 | -0.092 |
| gen_african | PDS_T1 | 1.037 | 0.310 | 3.342 | *** | 0.001 | 0.429 | 1.646 |
| gen_european | PDS_T1 | 0.913 | 0.368 | 2.480 |  | 0.013 | 0.191 | 1.635 |
| gen_east_asian | PDS_T1 | 0.371 | 0.164 | 2.259 |  | 0.024 | 0.049 | 0.692 |
| gen_american | PDS_T1 | 0.414 | 0.164 | 2.529 |  | 0.011 | 0.093 | 0.734 |
| age | dFA1 | 0.003 | 0.014 | 0.212 |  | 0.832 | -0.024 | 0.030 |
| BMI | dFA1 | -0.020 | 0.015 | -1.384 |  | 0.166 | -0.049 | 0.008 |
| SES | dFA1 | 0.044 | 0.016 | 2.690 | ** | 0.007 | 0.012 | 0.076 |
| gen_african | dFA1 | -0.341 | 0.321 | -1.062 |  | 0.288 | -0.971 | 0.289 |
| gen_european | dFA1 | -0.377 | 0.379 | -0.997 |  | 0.319 | -1.119 | 0.365 |
| gen_east_asian | dFA1 | -0.170 | 0.169 | -1.011 |  | 0.312 | -0.501 | 0.160 |
| gen_american | dFA1 | -0.190 | 0.170 | -1.119 |  | 0.263 | -0.523 | 0.143 |
| age | FA_T1 | 0.126 | 0.017 | 7.616 | *** | 0.000 | 0.094 | 0.159 |
| BMI | FA_T1 | 0.004 | 0.012 | 0.316 |  | 0.752 | -0.020 | 0.027 |
| SES | FA_T1 | 0.013 | 0.014 | 0.946 |  | 0.344 | -0.014 | 0.040 |
| gen_african | FA_T1 | -0.344 | 0.305 | -1.127 |  | 0.260 | -0.942 | 0.254 |
| gen_european | FA_T1 | -0.322 | 0.360 | -0.895 |  | 0.371 | -1.028 | 0.384 |
| gen_east_asian | FA_T1 | -0.162 | 0.160 | -1.011 |  | 0.312 | -0.475 | 0.152 |
| gen_american | FA_T1 | -0.137 | 0.161 | -0.850 |  | 0.395 | -0.452 | 0.178 |

| **Latent Factor Correlations** | | | | | | | |
| --- | --- | --- | --- | --- | --- | --- | --- |
| **Factor 1** | **Factor 2** | **r** | **sig** | **p** | **Lower.CI** | **Upper.CI** | **SE** |
| dPDS1 | dFA1 | 0.079 | *** | 0.000 | 0.036 | 0.122 | 0.022 |
| dPDS1 | dFA1 | 0.048 |  | 0.018 | 0.008 | 0.088 | 0.020 |

| **Latent Factor Variance/Residual Variance** | | | | | |
| --- | --- | --- | --- | --- | --- |
| **Factor 1** | **Factor 2** | **var** | **var.std** | **sig** | **p** |
| dPDS1 | dPDS1 | 0.538 | 0.696 | *** | 0 |
| dFA1 | dFA1 | 0.661 | 0.722 | *** | 0 |
| dPDS1 | dPDS1 | 0.447 | 0.724 | *** | 0 |
| dFA1 | dFA1 | 0.732 | 0.767 | *** | 0 |

**BLCS Mean Diffusivity**

Second, we run the model investigating white matter microstructural MD and pubertal maturation.

| **Model Significance** | | | |
| --- | --- | --- | --- |
| **Sample.Size** | **Chi.Square** | **df** | **p.value** |
| 9195 | 267.863 | 28 | 0 |

| **Model Fit Measures** | | | | | |
| --- | --- | --- | --- | --- | --- |
| **CFI** | **RMSEA** | **RMSEA.Lower** | **RMSEA.Upper** | **AIC** | **BIC** |
| 0.959 | 0.043 | 0.039 | 0.048 | 65663.04 | 66062.12 |

| **Covariances** | | | | | | | | | | |
| --- | --- | --- | --- | --- | --- | --- | --- | --- | --- | --- |
| lhs | op | rhs | group | label | est.std | se | z | pvalue | ci.lower | ci.upper |
| PDS_T1 | ~~ | MD_T1 | 1 |  | 0.01 | 0.02 | 0.53 | 0.60 | -.025 | .043 |
| dPDS1 | ~~ | dMD1 | 1 |  | -0.01 | 0.02 | -0.59 | 0.56 | -.060 | .032 |
| PDS_T1 | ~~ | MD_T1 | 2 |  | -0.00 | 0.01 | -0.30 | 0.76 | -.038 | .028 |
| dPDS1 | ~~ | dMD1 | 2 |  | -0.01 | 0.02 | -0.55 | 0.58 | -.049 | .027 |

| **Factor Loadings** | | | | | | | | |
| --- | --- | --- | --- | --- | --- | --- | --- | --- |
|  |  | **Standardized** | | | | | | |
| **Latent Factor** | **Indicator** | **Loadings** | **sig** | **p** | **Lower.CI** | **Upper.CI** | **SE** | **z** |
| dPDS1 | PDS_T2 | 0.940 | *** | 0 | 0.903 | 0.977 | 0.019 | 49.268 |
| dMD1 | MD_T2 | 0.894 | *** | 0 | 0.841 | 0.947 | 0.027 | 33.006 |
| dPDS1 | PDS_T2 | 0.979 | *** | 0 | 0.937 | 1.021 | 0.021 | 46.196 |
| dMD1 | MD_T2 | 0.910 | *** | 0 | 0.867 | 0.953 | 0.022 | 41.517 |

| **Regression Paths** | | | | | | | | |
| --- | --- | --- | --- | --- | --- | --- | --- | --- |
|  |  | **Standardized** | | | | | | |
| **Predictor** | **DV** | **Path Values** | **SE** | **z** | **sig** | **p** | **Lower.CI** | **Upper.CI** |
| PDS_T1 | PDS_T2 | 1.106 | 0.015 | 73.017 | *** | 0.000 | 1.077 | 1.136 |
| MD_T1 | MD_T2 | 1.009 | 0.019 | 52.211 | *** | 0.000 | 0.971 | 1.047 |
| MD_T1 | dPDS1 | -0.012 | 0.020 | -0.587 |  | 0.557 | -0.051 | 0.027 |
| PDS_T1 | dPDS1 | -0.586 | 0.014 | -41.983 | *** | 0.000 | -0.613 | -0.559 |
| PDS_T1 | dMD1 | -0.006 | 0.022 | -0.270 |  | 0.787 | -0.049 | 0.037 |
| MD_T1 | dMD1 | -0.449 | 0.023 | -19.425 | *** | 0.000 | -0.494 | -0.404 |
| age | dPDS1 | 0.073 | 0.012 | 6.185 | *** | 0.000 | 0.050 | 0.096 |
| BMI | dPDS1 | 0.101 | 0.013 | 7.851 | *** | 0.000 | 0.076 | 0.127 |
| SES | dPDS1 | 0.003 | 0.014 | 0.200 |  | 0.841 | -0.024 | 0.030 |
| gen_african | dPDS1 | -0.232 | 0.322 | -0.721 |  | 0.471 | -0.864 | 0.399 |
| gen_european | dPDS1 | -0.293 | 0.355 | -0.826 |  | 0.409 | -0.988 | 0.402 |
| gen_east_asian | dPDS1 | -0.074 | 0.105 | -0.709 |  | 0.478 | -0.279 | 0.131 |
| gen_american | dPDS1 | -0.094 | 0.143 | -0.657 |  | 0.511 | -0.374 | 0.186 |
| age | PDS_T1 | 0.137 | 0.013 | 10.469 | *** | 0.000 | 0.111 | 0.162 |
| BMI | PDS_T1 | 0.196 | 0.011 | 17.813 | *** | 0.000 | 0.175 | 0.218 |
| SES | PDS_T1 | -0.095 | 0.012 | -8.267 | *** | 0.000 | -0.118 | -0.073 |
| gen_african | PDS_T1 | 0.875 | 0.262 | 3.334 | *** | 0.001 | 0.361 | 1.389 |
| gen_european | PDS_T1 | 0.719 | 0.290 | 2.478 |  | 0.013 | 0.150 | 1.288 |
| gen_east_asian | PDS_T1 | 0.193 | 0.086 | 2.258 |  | 0.024 | 0.026 | 0.361 |
| gen_american | PDS_T1 | 0.293 | 0.116 | 2.527 |  | 0.011 | 0.066 | 0.521 |
| age | dMD1 | 0.032 | 0.016 | 2.029 |  | 0.042 | 0.001 | 0.064 |
| BMI | dMD1 | -0.003 | 0.015 | -0.195 |  | 0.845 | -0.033 | 0.027 |
| SES | dMD1 | -0.005 | 0.018 | -0.267 |  | 0.789 | -0.039 | 0.030 |
| gen_african | dMD1 | -0.025 | 0.391 | -0.064 |  | 0.949 | -0.792 | 0.742 |
| gen_european | dMD1 | -0.057 | 0.431 | -0.133 |  | 0.894 | -0.903 | 0.788 |
| gen_east_asian | dMD1 | -0.019 | 0.127 | -0.151 |  | 0.880 | -0.269 | 0.231 |
| gen_american | dMD1 | -0.065 | 0.173 | -0.373 |  | 0.709 | -0.404 | 0.275 |
| age | MD_T1 | -0.119 | 0.017 | -7.018 | *** | 0.000 | -0.152 | -0.086 |
| BMI | MD_T1 | -0.029 | 0.013 | -2.267 |  | 0.023 | -0.055 | -0.004 |
| SES | MD_T1 | 0.024 | 0.014 | 1.687 |  | 0.092 | -0.004 | 0.052 |
| gen_african | MD_T1 | 0.732 | 0.336 | 2.183 |  | 0.029 | 0.075 | 1.390 |
| gen_european | MD_T1 | 0.812 | 0.370 | 2.196 |  | 0.028 | 0.087 | 1.537 |
| gen_east_asian | MD_T1 | 0.228 | 0.109 | 2.092 |  | 0.036 | 0.014 | 0.441 |
| gen_american | MD_T1 | 0.325 | 0.149 | 2.187 |  | 0.029 | 0.034 | 0.617 |
| PDS_T1 | PDS_T2 | 1.027 | 0.019 | 53.422 | *** | 0.000 | 0.990 | 1.065 |
| MD_T1 | MD_T2 | 1.021 | 0.019 | 53.423 | *** | 0.000 | 0.983 | 1.058 |
| MD_T1 | dPDS1 | -0.018 | 0.017 | -1.028 |  | 0.304 | -0.052 | 0.016 |
| PDS_T1 | dPDS1 | -0.568 | 0.018 | -31.435 | *** | 0.000 | -0.603 | -0.532 |
| PDS_T1 | dMD1 | -0.018 | 0.023 | -0.768 |  | 0.442 | -0.063 | 0.027 |
| MD_T1 | dMD1 | -0.466 | 0.019 | -23.911 | *** | 0.000 | -0.504 | -0.427 |
| age | dPDS1 | 0.082 | 0.013 | 6.187 | *** | 0.000 | 0.056 | 0.108 |
| BMI | dPDS1 | 0.106 | 0.013 | 7.878 | *** | 0.000 | 0.080 | 0.132 |
| SES | dPDS1 | 0.003 | 0.015 | 0.201 |  | 0.841 | -0.027 | 0.033 |
| gen_african | dPDS1 | -0.246 | 0.341 | -0.721 |  | 0.471 | -0.914 | 0.422 |
| gen_european | dPDS1 | -0.332 | 0.402 | -0.827 |  | 0.408 | -1.120 | 0.455 |
| gen_east_asian | dPDS1 | -0.127 | 0.179 | -0.710 |  | 0.478 | -0.479 | 0.224 |
| gen_american | dPDS1 | -0.118 | 0.180 | -0.658 |  | 0.511 | -0.471 | 0.234 |
| age | PDS_T1 | 0.174 | 0.016 | 10.697 | *** | 0.000 | 0.142 | 0.206 |
| BMI | PDS_T1 | 0.230 | 0.012 | 18.407 | *** | 0.000 | 0.206 | 0.255 |
| SES | PDS_T1 | -0.120 | 0.014 | -8.439 | *** | 0.000 | -0.148 | -0.092 |
| gen_african | PDS_T1 | 1.039 | 0.311 | 3.346 | *** | 0.001 | 0.431 | 1.648 |
| gen_european | PDS_T1 | 0.915 | 0.368 | 2.484 |  | 0.013 | 0.193 | 1.637 |
| gen_east_asian | PDS_T1 | 0.372 | 0.164 | 2.263 |  | 0.024 | 0.050 | 0.694 |
| gen_american | PDS_T1 | 0.414 | 0.164 | 2.532 |  | 0.011 | 0.094 | 0.735 |
| age | dMD1 | 0.031 | 0.015 | 2.033 |  | 0.042 | 0.001 | 0.060 |
| BMI | dMD1 | -0.003 | 0.013 | -0.195 |  | 0.845 | -0.029 | 0.023 |
| SES | dMD1 | -0.004 | 0.017 | -0.267 |  | 0.790 | -0.037 | 0.028 |
| gen_african | dMD1 | -0.022 | 0.346 | -0.064 |  | 0.949 | -0.699 | 0.655 |
| gen_european | dMD1 | -0.054 | 0.408 | -0.133 |  | 0.894 | -0.853 | 0.745 |
| gen_east_asian | dMD1 | -0.028 | 0.182 | -0.151 |  | 0.880 | -0.385 | 0.330 |
| gen_american | dMD1 | -0.068 | 0.182 | -0.373 |  | 0.709 | -0.424 | 0.288 |
| age | MD_T1 | -0.113 | 0.016 | -6.972 | *** | 0.000 | -0.145 | -0.081 |
| BMI | MD_T1 | -0.026 | 0.011 | -2.265 |  | 0.023 | -0.048 | -0.003 |
| SES | MD_T1 | 0.023 | 0.014 | 1.685 |  | 0.092 | -0.004 | 0.049 |
| gen_african | MD_T1 | 0.651 | 0.299 | 2.179 |  | 0.029 | 0.066 | 1.236 |
| gen_european | MD_T1 | 0.773 | 0.353 | 2.193 |  | 0.028 | 0.082 | 1.464 |
| gen_east_asian | MD_T1 | 0.328 | 0.157 | 2.088 |  | 0.037 | 0.020 | 0.636 |
| gen_american | MD_T1 | 0.344 | 0.158 | 2.183 |  | 0.029 | 0.035 | 0.653 |

| **Latent Factor Correlations** | | | | | | | |
| --- | --- | --- | --- | --- | --- | --- | --- |
| **Factor 1** | **Factor 2** | **r** | **sig** | **p** | **Lower.CI** | **Upper.CI** | **SE** |
| dPDS1 | dMD1 | -0.014 |  | 0.558 | -0.060 | 0.032 | 0.023 |
| dPDS1 | dMD1 | -0.011 |  | 0.584 | -0.049 | 0.027 | 0.019 |

| **Latent Factor Variance/Residual Variance** | | | | | |
| --- | --- | --- | --- | --- | --- |
| **Factor 1** | **Factor 2** | **var** | **var.std** | **sig** | **p** |
| dPDS1 | dPDS1 | 0.537 | 0.697 | *** | 0 |
| dMD1 | dMD1 | 0.581 | 0.791 | *** | 0 |
| dPDS1 | dPDS1 | 0.447 | 0.724 | *** | 0 |
| dMD1 | dMD1 | 0.657 | 0.777 | *** | 0 |

**BLCS Cortical Thickness**

Third, we run the model testing cortical thickness and pubertal maturation:

| **Model Significance** | | | |
| --- | --- | --- | --- |
| **Sample.Size** | **Chi.Square** | **df** | **p.value** |
| 9195 | 274.47 | 28 | 0 |

| **Model Fit Measures** | | | | | |
| --- | --- | --- | --- | --- | --- |
| **CFI** | **RMSEA** | **RMSEA.Lower** | **RMSEA.Upper** | **AIC** | **BIC** |
| 0.963 | 0.044 | 0.039 | 0.049 | 66842.73 | 67241.81 |

| **Covariances** | | | | | | | | | | |
| --- | --- | --- | --- | --- | --- | --- | --- | --- | --- | --- |
| lhs | op | rhs | group | label | est.std | se | z | pvalue | ci.lower | ci.upper |
| PDS_T1 | ~~ | Thickness_T1 | 1 |  | -0.01 | 0.02 | -0.71 | 0.48 | -.045 | .021 |
| dPDS1 | ~~ | dThickness1 | 1 |  | -0.03 | 0.01 | -2.68 | 0.00 | -.101 | -.016 |
| PDS_T1 | ~~ | Thickness_T1 | 2 |  | 0.00 | 0.01 | 0.05 | 0.96 | -.029 | .031 |
| dPDS1 | ~~ | dThickness1 | 2 |  | -0.00 | 0.01 | -0.07 | 0.94 | -.038 | .036 |

| **Factor Loadings** | | | | | | | | |
| --- | --- | --- | --- | --- | --- | --- | --- | --- |
|  |  | **Standardized** | | | | | | |
| **Latent Factor** | **Indicator** | **Loadings** | **sig** | **p** | **Lower.CI** | **Upper.CI** | **SE** | **z** |
| dPDS1 | PDS_T2 | 0.939 | *** | 0 | 0.902 | 0.977 | 0.019 | 49.292 |
| dThickness1 | Thickness_T2 | 0.845 | *** | 0 | 0.797 | 0.894 | 0.025 | 34.271 |
| dPDS1 | PDS_T2 | 0.979 | *** | 0 | 0.937 | 1.020 | 0.021 | 46.194 |
| dThickness1 | Thickness_T2 | 0.873 | *** | 0 | 0.831 | 0.916 | 0.022 | 40.400 |

| **Regression Paths** | | | | | | | | |
| --- | --- | --- | --- | --- | --- | --- | --- | --- |
|  |  | **Standardized** | | | | | | |
| **Predictor** | **DV** | **Path Values** | **SE** | **z** | **sig** | **p** | **Lower.CI** | **Upper.CI** |
| PDS_T1 | PDS_T2 | 1.107 | 0.015 | 72.983 | *** | 0.000 | 1.077 | 1.136 |
| Thickness_T1 | Thickness_T2 | 1.022 | 0.019 | 54.917 | *** | 0.000 | 0.986 | 1.059 |
| Thickness_T1 | dPDS1 | 0.012 | 0.019 | 0.623 |  | 0.533 | -0.025 | 0.048 |
| PDS_T1 | dPDS1 | -0.584 | 0.014 | -41.535 | *** | 0.000 | -0.612 | -0.557 |
| PDS_T1 | dThickness1 | -0.105 | 0.023 | -4.526 | *** | 0.000 | -0.151 | -0.060 |
| Thickness_T1 | dThickness1 | -0.463 | 0.020 | -22.700 | *** | 0.000 | -0.503 | -0.423 |
| age | dPDS1 | 0.073 | 0.012 | 6.297 | *** | 0.000 | 0.051 | 0.096 |
| BMI | dPDS1 | 0.101 | 0.013 | 7.831 | *** | 0.000 | 0.076 | 0.126 |
| SES | dPDS1 | 0.002 | 0.014 | 0.142 |  | 0.887 | -0.025 | 0.029 |
| gen_african | dPDS1 | -0.218 | 0.322 | -0.677 |  | 0.499 | -0.850 | 0.414 |
| gen_european | dPDS1 | -0.276 | 0.355 | -0.778 |  | 0.437 | -0.972 | 0.420 |
| gen_east_asian | dPDS1 | -0.069 | 0.105 | -0.661 |  | 0.509 | -0.275 | 0.136 |
| gen_american | dPDS1 | -0.087 | 0.143 | -0.611 |  | 0.541 | -0.368 | 0.193 |
| age | PDS_T1 | 0.136 | 0.013 | 10.469 | *** | 0.000 | 0.111 | 0.162 |
| BMI | PDS_T1 | 0.196 | 0.011 | 17.831 | *** | 0.000 | 0.175 | 0.218 |
| SES | PDS_T1 | -0.095 | 0.012 | -8.251 | *** | 0.000 | -0.118 | -0.072 |
| gen_african | PDS_T1 | 0.876 | 0.262 | 3.338 | *** | 0.001 | 0.362 | 1.390 |
| gen_european | PDS_T1 | 0.720 | 0.290 | 2.482 |  | 0.013 | 0.151 | 1.289 |
| gen_east_asian | PDS_T1 | 0.193 | 0.086 | 2.262 |  | 0.024 | 0.026 | 0.361 |
| gen_american | PDS_T1 | 0.294 | 0.116 | 2.530 |  | 0.011 | 0.066 | 0.521 |
| age | dThickness1 | -0.005 | 0.016 | -0.305 |  | 0.760 | -0.035 | 0.026 |
| BMI | dThickness1 | 0.002 | 0.016 | 0.148 |  | 0.882 | -0.029 | 0.033 |
| SES | dThickness1 | 0.033 | 0.017 | 1.888 |  | 0.059 | -0.001 | 0.066 |
| gen_african | dThickness1 | -0.521 | 0.392 | -1.328 |  | 0.184 | -1.291 | 0.248 |
| gen_european | dThickness1 | -0.525 | 0.433 | -1.213 |  | 0.225 | -1.372 | 0.323 |
| gen_east_asian | dThickness1 | -0.176 | 0.127 | -1.380 |  | 0.167 | -0.425 | 0.074 |
| gen_american | dThickness1 | -0.239 | 0.174 | -1.377 |  | 0.169 | -0.579 | 0.101 |
| age | Thickness_T1 | -0.070 | 0.016 | -4.482 | *** | 0.000 | -0.101 | -0.040 |
| BMI | Thickness_T1 | -0.063 | 0.013 | -5.000 | *** | 0.000 | -0.087 | -0.038 |
| SES | Thickness_T1 | 0.022 | 0.013 | 1.726 |  | 0.084 | -0.003 | 0.047 |
| gen_african | Thickness_T1 | -0.050 | 0.313 | -0.160 |  | 0.873 | -0.663 | 0.563 |
| gen_european | Thickness_T1 | 0.101 | 0.345 | 0.292 |  | 0.770 | -0.575 | 0.776 |
| gen_east_asian | Thickness_T1 | -0.023 | 0.102 | -0.231 |  | 0.818 | -0.223 | 0.176 |
| gen_american | Thickness_T1 | -0.023 | 0.138 | -0.164 |  | 0.870 | -0.294 | 0.249 |
| PDS_T1 | PDS_T2 | 1.027 | 0.019 | 53.433 | *** | 0.000 | 0.989 | 1.065 |
| Thickness_T1 | Thickness_T2 | 0.992 | 0.017 | 58.371 | *** | 0.000 | 0.959 | 1.025 |
| Thickness_T1 | dPDS1 | -0.024 | 0.018 | -1.395 |  | 0.163 | -0.059 | 0.010 |
| PDS_T1 | dPDS1 | -0.568 | 0.018 | -31.481 | *** | 0.000 | -0.604 | -0.533 |
| PDS_T1 | dThickness1 | -0.002 | 0.022 | -0.098 |  | 0.922 | -0.045 | 0.040 |
| Thickness_T1 | dThickness1 | -0.444 | 0.019 | -23.978 | *** | 0.000 | -0.481 | -0.408 |
| age | dPDS1 | 0.083 | 0.013 | 6.297 | *** | 0.000 | 0.057 | 0.109 |
| BMI | dPDS1 | 0.106 | 0.013 | 7.856 | *** | 0.000 | 0.079 | 0.132 |
| SES | dPDS1 | 0.002 | 0.015 | 0.142 |  | 0.887 | -0.028 | 0.032 |
| gen_african | dPDS1 | -0.231 | 0.341 | -0.677 |  | 0.498 | -0.899 | 0.438 |
| gen_european | dPDS1 | -0.313 | 0.402 | -0.778 |  | 0.436 | -1.101 | 0.475 |
| gen_east_asian | dPDS1 | -0.119 | 0.179 | -0.661 |  | 0.509 | -0.470 | 0.233 |
| gen_american | dPDS1 | -0.110 | 0.180 | -0.612 |  | 0.541 | -0.463 | 0.243 |
| age | PDS_T1 | 0.174 | 0.016 | 10.694 | *** | 0.000 | 0.142 | 0.206 |
| BMI | PDS_T1 | 0.230 | 0.012 | 18.424 | *** | 0.000 | 0.206 | 0.255 |
| SES | PDS_T1 | -0.120 | 0.014 | -8.420 | *** | 0.000 | -0.148 | -0.092 |
| gen_african | PDS_T1 | 1.041 | 0.311 | 3.350 | *** | 0.001 | 0.432 | 1.649 |
| gen_european | PDS_T1 | 0.917 | 0.368 | 2.488 |  | 0.013 | 0.194 | 1.639 |
| gen_east_asian | PDS_T1 | 0.372 | 0.164 | 2.267 |  | 0.023 | 0.050 | 0.694 |
| gen_american | PDS_T1 | 0.415 | 0.164 | 2.535 |  | 0.011 | 0.094 | 0.736 |
| age | dThickness1 | -0.005 | 0.015 | -0.305 |  | 0.760 | -0.034 | 0.025 |
| BMI | dThickness1 | 0.002 | 0.014 | 0.148 |  | 0.882 | -0.025 | 0.029 |
| SES | dThickness1 | 0.031 | 0.016 | 1.885 |  | 0.059 | -0.001 | 0.063 |
| gen_african | dThickness1 | -0.463 | 0.349 | -1.324 |  | 0.185 | -1.148 | 0.222 |
| gen_european | dThickness1 | -0.499 | 0.412 | -1.210 |  | 0.226 | -1.307 | 0.309 |
| gen_east_asian | dThickness1 | -0.253 | 0.184 | -1.377 |  | 0.169 | -0.613 | 0.107 |
| gen_american | dThickness1 | -0.252 | 0.184 | -1.373 |  | 0.170 | -0.613 | 0.108 |
| age | Thickness_T1 | -0.071 | 0.016 | -4.484 | *** | 0.000 | -0.102 | -0.040 |
| BMI | Thickness_T1 | -0.059 | 0.012 | -5.000 | *** | 0.000 | -0.082 | -0.036 |
| SES | Thickness_T1 | 0.022 | 0.013 | 1.726 |  | 0.084 | -0.003 | 0.047 |
| gen_african | Thickness_T1 | -0.047 | 0.296 | -0.160 |  | 0.873 | -0.627 | 0.532 |
| gen_european | Thickness_T1 | 0.102 | 0.349 | 0.292 |  | 0.770 | -0.582 | 0.785 |
| gen_east_asian | Thickness_T1 | -0.036 | 0.156 | -0.231 |  | 0.818 | -0.341 | 0.269 |
| gen_american | Thickness_T1 | -0.025 | 0.156 | -0.164 |  | 0.870 | -0.331 | 0.280 |

| **Latent Factor Correlations** | | | | | | | |
| --- | --- | --- | --- | --- | --- | --- | --- |
| **Factor 1** | **Factor 2** | **r** | **sig** | **p** | **Lower.CI** | **Upper.CI** | **SE** |
| dPDS1 | dThickness1 | -0.058 | ** | 0.008 | -0.101 | -0.016 | 0.022 |
| dPDS1 | dThickness1 | -0.001 |  | 0.942 | -0.038 | 0.036 | 0.019 |

| **Latent Factor Variance/Residual Variance** | | | | | |
| --- | --- | --- | --- | --- | --- |
| **Factor 1** | **Factor 2** | **var** | **var.std** | **sig** | **p** |
| dPDS1 | dPDS1 | 0.537 | 0.698 | *** | 0 |
| dThickness1 | dThickness1 | 0.534 | 0.786 | *** | 0 |
| dPDS1 | dPDS1 | 0.447 | 0.724 | *** | 0 |
| dThickness1 | dThickness1 | 0.626 | 0.809 | *** | 0 |

**BLCS Surface Area**

Lastly, we run the model testing surface area and pubertal maturation:

| **Model Significance** | | | |
| --- | --- | --- | --- |
| **Sample.Size** | **Chi.Square** | **df** | **p.value** |
| 9195 | 275.743 | 28 | 0 |

| **Model Fit Measures** | | | | | |
| --- | --- | --- | --- | --- | --- |
| **CFI** | **RMSEA** | **RMSEA.Lower** | **RMSEA.Upper** | **AIC** | **BIC** |
| 0.982 | 0.044 | 0.039 | 0.049 | 56596.72 | 56995.8 |

| **Covariances** | | | | | | | | | | |
| --- | --- | --- | --- | --- | --- | --- | --- | --- | --- | --- |
| lhs | op | rhs | group | label | est.std | se | z | pvalue | ci.lower | ci.upper |
| PDS_T1 | ~~ | Area_T1 | 1 |  | -0.00 | 0.01 | -0.04 | 0.97 | -.031 | .030 |
| dPDS1 | ~~ | dArea1 | 1 |  | 0.00 | 0.01 | 0.11 | 0.91 | -.040 | .045 |
| PDS_T1 | ~~ | Area_T1 | 2 |  | 0.04 | 0.01 | 3.64 | 0.00 | .026 | .084 |
| dPDS1 | ~~ | dArea1 | 2 |  | 0.02 | 0.00 | 3.95 | 0.00 | .042 | .125 |

| **Factor Loadings** | | | | | | | | |
| --- | --- | --- | --- | --- | --- | --- | --- | --- |
|  |  | **Standardized** | | | | | | |
| **Latent Factor** | **Indicator** | **Loadings** | **sig** | **p** | **Lower.CI** | **Upper.CI** | **SE** | **z** |
| dPDS1 | PDS_T2 | 0.940 | *** | 0 | 0.903 | 0.978 | 0.019 | 49.233 |
| dArea1 | Area_T2 | 0.392 | *** | 0 | 0.357 | 0.428 | 0.018 | 21.449 |
| dPDS1 | PDS_T2 | 0.980 | *** | 0 | 0.938 | 1.021 | 0.021 | 46.123 |
| dArea1 | Area_T2 | 0.353 | *** | 0 | 0.329 | 0.377 | 0.012 | 28.441 |

| **Regression Paths** | | | | | | | | |
| --- | --- | --- | --- | --- | --- | --- | --- | --- |
|  |  | **Standardized** | | | | | | |
| Predictor | DV | Path Values | SE | z | sig | p | Lower.CI | Upper.CI |
| PDS_T1 | PDS_T2 | 1.106 | 0.015 | 72.988 | *** | 0.000 | 1.077 | 1.136 |
| Area_T1 | Area_T2 | 0.994 | 0.008 | 125.638 | *** | 0.000 | 0.979 | 1.010 |
| Area_T1 | dPDS1 | 0.006 | 0.019 | 0.313 |  | 0.754 | -0.031 | 0.043 |
| PDS_T1 | dPDS1 | -0.585 | 0.014 | -41.777 | *** | 0.000 | -0.613 | -0.558 |
| PDS_T1 | dArea1 | -0.210 | 0.023 | -9.258 | *** | 0.000 | -0.255 | -0.166 |
| Area_T1 | dArea1 | -0.216 | 0.023 | -9.341 | *** | 0.000 | -0.262 | -0.171 |
| age | dPDS1 | 0.075 | 0.012 | 6.427 | *** | 0.000 | 0.052 | 0.097 |
| BMI | dPDS1 | 0.103 | 0.013 | 7.999 | *** | 0.000 | 0.078 | 0.129 |
| SES | dPDS1 | 0.002 | 0.014 | 0.147 |  | 0.883 | -0.025 | 0.029 |
| gen_african | dPDS1 | -0.229 | 0.322 | -0.712 |  | 0.476 | -0.860 | 0.402 |
| gen_european | dPDS1 | -0.287 | 0.355 | -0.809 |  | 0.419 | -0.981 | 0.408 |
| gen_east_asian | dPDS1 | -0.073 | 0.105 | -0.696 |  | 0.487 | -0.278 | 0.132 |
| gen_american | dPDS1 | -0.094 | 0.143 | -0.656 |  | 0.512 | -0.373 | 0.186 |
| age | PDS_T1 | 0.136 | 0.013 | 10.465 | *** | 0.000 | 0.111 | 0.162 |
| BMI | PDS_T1 | 0.196 | 0.011 | 17.799 | *** | 0.000 | 0.174 | 0.217 |
| SES | PDS_T1 | -0.095 | 0.012 | -8.216 | *** | 0.000 | -0.117 | -0.072 |
| gen_african | PDS_T1 | 0.856 | 0.262 | 3.269 | ** | 0.001 | 0.343 | 1.370 |
| gen_european | PDS_T1 | 0.698 | 0.290 | 2.409 |  | 0.016 | 0.130 | 1.266 |
| gen_east_asian | PDS_T1 | 0.187 | 0.085 | 2.187 |  | 0.029 | 0.019 | 0.354 |
| gen_american | PDS_T1 | 0.285 | 0.116 | 2.464 |  | 0.014 | 0.058 | 0.512 |
| age | dArea1 | -0.154 | 0.022 | -7.121 | *** | 0.000 | -0.196 | -0.111 |
| BMI | dArea1 | -0.052 | 0.016 | -3.256 | ** | 0.001 | -0.084 | -0.021 |
| SES | dArea1 | -0.005 | 0.017 | -0.266 |  | 0.790 | -0.038 | 0.029 |
| gen_african | dArea1 | -0.272 | 0.405 | -0.671 |  | 0.502 | -1.066 | 0.523 |
| gen_european | dArea1 | -0.217 | 0.446 | -0.487 |  | 0.626 | -1.090 | 0.656 |
| gen_east_asian | dArea1 | -0.073 | 0.131 | -0.554 |  | 0.580 | -0.330 | 0.185 |
| gen_american | dArea1 | -0.117 | 0.179 | -0.654 |  | 0.513 | -0.468 | 0.234 |
| age | Area_T1 | 0.012 | 0.012 | 0.940 |  | 0.347 | -0.013 | 0.036 |
| BMI | Area_T1 | 0.067 | 0.012 | 5.707 | *** | 0.000 | 0.044 | 0.090 |
| SES | Area_T1 | 0.106 | 0.013 | 8.393 | *** | 0.000 | 0.081 | 0.131 |
| gen_african | Area_T1 | -0.064 | 0.304 | -0.209 |  | 0.834 | -0.659 | 0.532 |
| gen_european | Area_T1 | 0.189 | 0.335 | 0.564 |  | 0.573 | -0.467 | 0.845 |
| gen_east_asian | Area_T1 | 0.046 | 0.098 | 0.466 |  | 0.641 | -0.147 | 0.239 |
| gen_american | Area_T1 | -0.006 | 0.135 | -0.047 |  | 0.963 | -0.270 | 0.257 |
| PDS_T1 | PDS_T2 | 1.027 | 0.019 | 53.380 | *** | 0.000 | 0.990 | 1.065 |
| Area_T1 | Area_T2 | 1.007 | 0.007 | 141.086 | *** | 0.000 | 0.993 | 1.021 |
| Area_T1 | dPDS1 | -0.021 | 0.017 | -1.256 |  | 0.209 | -0.055 | 0.012 |
| PDS_T1 | dPDS1 | -0.568 | 0.018 | -31.481 | *** | 0.000 | -0.604 | -0.533 |
| PDS_T1 | dArea1 | -0.078 | 0.021 | -3.642 | *** | 0.000 | -0.119 | -0.036 |
| Area_T1 | dArea1 | -0.212 | 0.021 | -10.313 | *** | 0.000 | -0.253 | -0.172 |
| age | dPDS1 | 0.085 | 0.013 | 6.426 | *** | 0.000 | 0.059 | 0.110 |
| BMI | dPDS1 | 0.108 | 0.013 | 8.022 | *** | 0.000 | 0.082 | 0.135 |
| SES | dPDS1 | 0.002 | 0.015 | 0.147 |  | 0.883 | -0.028 | 0.033 |
| gen_african | dPDS1 | -0.243 | 0.341 | -0.712 |  | 0.476 | -0.911 | 0.425 |
| gen_european | dPDS1 | -0.325 | 0.402 | -0.809 |  | 0.419 | -1.113 | 0.463 |
| gen_east_asian | dPDS1 | -0.125 | 0.179 | -0.696 |  | 0.487 | -0.476 | 0.227 |
| gen_american | dPDS1 | -0.118 | 0.180 | -0.656 |  | 0.512 | -0.471 | 0.234 |
| age | PDS_T1 | 0.173 | 0.016 | 10.693 | *** | 0.000 | 0.142 | 0.205 |
| BMI | PDS_T1 | 0.230 | 0.012 | 18.387 | *** | 0.000 | 0.205 | 0.254 |
| SES | PDS_T1 | -0.120 | 0.014 | -8.385 | *** | 0.000 | -0.148 | -0.092 |
| gen_african | PDS_T1 | 1.017 | 0.310 | 3.281 | ** | 0.001 | 0.410 | 1.625 |
| gen_european | PDS_T1 | 0.888 | 0.368 | 2.414 |  | 0.016 | 0.167 | 1.609 |
| gen_east_asian | PDS_T1 | 0.359 | 0.164 | 2.192 |  | 0.028 | 0.038 | 0.681 |
| gen_american | PDS_T1 | 0.403 | 0.163 | 2.469 |  | 0.014 | 0.083 | 0.724 |
| age | dArea1 | -0.163 | 0.021 | -7.603 | *** | 0.000 | -0.205 | -0.121 |
| BMI | dArea1 | -0.051 | 0.016 | -3.252 | ** | 0.001 | -0.082 | -0.020 |
| SES | dArea1 | -0.005 | 0.018 | -0.266 |  | 0.790 | -0.040 | 0.031 |
| gen_african | dArea1 | -0.270 | 0.400 | -0.674 |  | 0.500 | -1.054 | 0.514 |
| gen_european | dArea1 | -0.231 | 0.472 | -0.489 |  | 0.625 | -1.155 | 0.694 |
| gen_east_asian | dArea1 | -0.117 | 0.210 | -0.557 |  | 0.578 | -0.529 | 0.295 |
| gen_american | dArea1 | -0.138 | 0.210 | -0.657 |  | 0.511 | -0.550 | 0.274 |
| age | Area_T1 | 0.011 | 0.012 | 0.940 |  | 0.347 | -0.012 | 0.034 |
| BMI | Area_T1 | 0.058 | 0.010 | 5.710 | *** | 0.000 | 0.038 | 0.078 |
| SES | Area_T1 | 0.099 | 0.012 | 8.409 | *** | 0.000 | 0.076 | 0.123 |
| gen_african | Area_T1 | -0.056 | 0.267 | -0.209 |  | 0.834 | -0.580 | 0.468 |
| gen_european | Area_T1 | 0.178 | 0.316 | 0.564 |  | 0.573 | -0.441 | 0.797 |
| gen_east_asian | Area_T1 | 0.065 | 0.140 | 0.466 |  | 0.641 | -0.210 | 0.341 |
| gen_american | Area_T1 | -0.007 | 0.141 | -0.047 |  | 0.963 | -0.283 | 0.270 |

| **Latent Factor Correlations** | | | | | | | |
| --- | --- | --- | --- | --- | --- | --- | --- |
| **Factor 1** | **Factor 2** | **r** | **sig** | **p** | **Lower.CI** | **Upper.CI** | **SE** |
| dPDS1 | dArea1 | 0.002 |  | 0.913 | -0.040 | 0.045 | 0.022 |
| dPDS1 | dArea1 | 0.083 | *** | 0.000 | 0.042 | 0.125 | 0.021 |

| **Latent Factor Variance/Residual Variance** | | | | | |
| --- | --- | --- | --- | --- | --- |
| **Factor 1** | **Factor 2** | **var** | **var.std** | **sig** | **p** |
| dPDS1 | dPDS1 | 0.538 | 0.697 | *** | 0 |
| dArea1 | dArea1 | 0.098 | 0.860 | *** | 0 |
| dPDS1 | dPDS1 | 0.447 | 0.724 | *** | 0 |
| dArea1 | dArea1 | 0.094 | 0.906 | *** | 0 |

**Group effects - Area regressions**

We also want to look at some tests where we look at group effects by fixing model parameters (regression coefficients and covariance) and test sex differences with ANOVAs.

Let’s do this using Area data first.

Reg 1: dArea1 ~ c(e,e)*PDS_T1

Reg 2: dPDS1 ~ c(e,e)*Area_T1

**Comparing group effects - Area**

Let’s compare the models by running separate ANOVAs, each testing the original Area BLCS model against the new group effects regression models.

|  | **Df** | **AIC** | **BIC** | **Chisq** | **Chisq diff** | **Df diff** | **Pr(>Chisq)** |
| --- | --- | --- | --- | --- | --- | --- | --- |
| fitBLCS_Area | 28 | 56596.72 | 56995.80 | 275.7433 | NA | NA | NA |
| fitBLCS_Area_reg_1_ | 29 | 56607.24 | 56999.19 | 288.2602 | 12.25914 | 1 | 0.000463 |

|  | **Df** | **AIC** | **BIC** | **Chisq** | **Chisq diff** | **Df diff** | **Pr(>Chisq)** |
| --- | --- | --- | --- | --- | --- | --- | --- |
| fitBLCS_Area | 28 | 56596.72 | 56995.80 | 275.7433 | NA | NA | NA |
| fitBLCS_Area_reg_2_ | 29 | 56595.80 | 56987.76 | 276.8277 | 1.031558 | 1 | 0.3097929 |

**Group effects - Thickness regressions**

Now, let’s re-run the ANOVAs on Thickness measures.

Reg 1: dThickness1 ~ c(e,e)*PDS_T1

Reg 2: dPDS1 ~ c(e,e)*Thickness_T1

**Comparing group effects - Thickness**

Let’s compare the models by running separate ANOVAs, each testing the original Thickness BLCS model against the new group effects regression models.

|  | **Df** | **AIC** | **BIC** | **Chisq** | **Chisq diff** | **Df diff** | **Pr(>Chisq)** |
| --- | --- | --- | --- | --- | --- | --- | --- |
| fitBLCS_Thickness | 28 | 66842.73 | 67241.81 | 274.4699 | NA | NA | NA |
| fitBLCS_Thickness_reg_1_ | 29 | 66850.52 | 67242.47 | 284.2562 | 8.747091 | 1 | 0.003101 |

|  | **Df** | **AIC** | **BIC** | **Chisq** | **Chisq diff** | **Df diff** | **Pr(>Chisq)** |
| --- | --- | --- | --- | --- | --- | --- | --- |
| fitBLCS_Thickness | 28 | 66842.73 | 67241.81 | 274.4699 | NA | NA | NA |
| fitBLCS_Thickness_reg_2_ | 29 | 66842.67 | 67234.62 | 276.4068 | 1.95036 | 1 | 0.1625481 |

**Group effects - MD regressions**

Now, let’s re-run the ANOVAs on MD measures.

Reg 1: dMD1 ~ c(e,e)*PDS_T1

Reg 2: dPDS1 ~ c(e,e)*MD_T1

**Comparing group effects - MD**

Let’s compare the models by running separate ANOVAs, each testing the original MD BLCS model against the new group effects regression models.

|  | **Df** | **AIC** | **BIC** | **Chisq** | **Chisq diff** | **Df diff** | **Pr(>Chisq)** |
| --- | --- | --- | --- | --- | --- | --- | --- |
| fitBLCS_MD | 28 | 65663.05 | 66062.12 | 267.8634 | NA | NA | NA |
| fitBLCS_MD_reg_1_ | 29 | 65661.32 | 66053.27 | 268.1390 | 0.2478404 | 1 | 0.6185999 |

|  | **Df** | **AIC** | **BIC** | **Chisq** | **Chisq diff** | **Df diff** | **Pr(>Chisq)** |
| --- | --- | --- | --- | --- | --- | --- | --- |
| fitBLCS_MD | 28 | 65663.05 | 66062.12 | 267.8634 | NA | NA | NA |
| fitBLCS_MD_reg_2_ | 29 | 65661.06 | 66053.02 | 267.8821 | 0.0177922 | 1 | 0.8938869 |

**Group effects - FA regressions**

Now, let’s re-run the ANOVAs on FA measures.

Reg 1: dFA1 ~ c(e,e)*PDS_T1

Reg 2: dPDS1 ~ c(e,e)*FA_T1

**Comparing group effects - FA**

Let’s compare the models by running separate ANOVAs, each testing the original FA BLCS model against the new group effects regression models.

|  | **Df** | **AIC** | **BIC** | **Chisq** | **Chisq diff** | **Df diff** | **Pr(>Chisq)** |
| --- | --- | --- | --- | --- | --- | --- | --- |
| fitBLCS_FA | 28 | 66202.68 | 66601.76 | 272.4489 | NA | NA | NA |
| fitBLCS_FA_reg_1_ | 29 | 66200.68 | 66592.64 | 272.4504 | 0.0015748 | 1 | 0.9683452 |

|  | **Df** | **AIC** | **BIC** | **Chisq** | **Chisq diff** | **Df diff** | **Pr(>Chisq)** |
| --- | --- | --- | --- | --- | --- | --- | --- |
| fitBLCS_FA | 28 | 66202.68 | 66601.76 | 272.4489 | NA | NA | NA |
| fitBLCS_FA_reg_2_ | 29 | 66201.69 | 66593.65 | 273.4592 | 0.9510997 | 1 | 0.3294395 |

**Group effects - Area correlation**

The next section we look at some tests where we look at group effects by fixing covariance parameters test sex differences with ANOVAs. Let’s do this with Area data first.

Corr 1: PDS_T1 ~~ c(e,e)*Area_T1

Corr 2: dPDS1 ~~ c(e,e)*dArea1

**Comparing group effects II - Area**

Let’s compare the models by running separate ANOVAs, each testing the original Area BLCS model against the two new group effects models.

|  | **Df** | **AIC** | **BIC** | **Chisq** | **Chisq diff** | **Df diff** | **Pr(>Chisq)** |
| --- | --- | --- | --- | --- | --- | --- | --- |
| fitBLCS_Area | 28 | 56596.72 | 56995.80 | 275.7433 | NA | NA | NA |
| fitBLCS_Area_corr_1_ | 29 | 56599.65 | 56991.61 | 280.6760 | 5.386443 | 1 | 0.0202938 |

|  | **Df** | **AIC** | **BIC** | **Chisq** | **Chisq diff** | **Df diff** | **Pr(>Chisq)** |
| --- | --- | --- | --- | --- | --- | --- | --- |
| fitBLCS_Area | 28 | 56596.72 | 56995.80 | 275.7433 | NA | NA | NA |
| fitBLCS_Area_corr_2_ | 29 | 56600.57 | 56992.52 | 281.5891 | 6.139957 | 1 | 0.0132161 |

**Group effects - Thickness correlation**

Corr 1: PDS_T1 ~~ c(e,e)*Thickness_T1

Corr 2: dPDS1 ~~ c(e,e)*dThickness1

**Comparing group effects II – Thickness**

Let’s compare the models by running separate ANOVAs, each testing the original Thickness BLCS model against the two new group effects models.

|  | **Df** | **AIC** | **BIC** | **Chisq** | **Chisq diff** | **Df diff** | **Pr(>Chisq)** |
| --- | --- | --- | --- | --- | --- | --- | --- |
| fitBLCS_Thickness | 28 | 66842.73 | 67241.81 | 274.4699 | NA | NA | NA |
| fitBLCS_Thickness_corr_1_ | 29 | 66841.10 | 67233.05 | 274.8386 | 0.3698796 | 1 | 0.5430699 |

|  | **Df** | **AIC** | **BIC** | **Chisq** | **Chisq diff** | **Df diff** | **Pr(>Chisq)** |
| --- | --- | --- | --- | --- | --- | --- | --- |
| fitBLCS_Thickness | 28 | 66842.73 | 67241.81 | 274.4699 | NA | NA | NA |
| fitBLCS_Thickness_corr_2_ | 29 | 66844.49 | 67236.44 | 278.2276 | 3.862544 | 1 | 0.0493754 |

**Group effects - MD correlation**

Corr 1: PDS_T1 ~~ c(e,e)*MD_T1

Corr 2: dPDS1 ~~ c(e,e)*dMD1

**Comparing group effects II – MD**

Let’s compare the models by running separate ANOVAs, each testing the original MD BLCS model against the two new group effects models.

|  | **Df** | **AIC** | **BIC** | **Chisq** | **Chisq diff** | **Df diff** | **Pr(>Chisq)** |
| --- | --- | --- | --- | --- | --- | --- | --- |
| fitBLCS_MD | 28 | 65663.05 | 66062.12 | 267.8634 | NA | NA | NA |
| fitBLCS_MD_corr_1_ | 29 | 65661.42 | 66053.37 | 268.2368 | 0.3606937 | 1 | 0.5481212 |

|  | **Df** | **AIC** | **BIC** | **Chisq** | **Chisq diff** | **Df diff** | **Pr(>Chisq)** |
| --- | --- | --- | --- | --- | --- | --- | --- |
| fitBLCS_MD | 28 | 65663.05 | 66062.12 | 267.8634 | NA | NA | NA |
| fitBLCS_MD_corr_2_ | 29 | 65661.06 | 66053.01 | 267.8765 | 0.0130719 | 1 | 0.9089743 |

**Group effects - FA correlation**

Corr 1: PDS_T1 ~~ c(e,e)*FA_T1

Corr 2: dPDS1 ~~ c(e,e)*dFA1

**Comparing group effects II - FA**

Let’s compare the models by running separate ANOVAs, each testing the original FA BLCS model against the two new group effects models.

|  | **Df** | **AIC** | **BIC** | **Chisq** | **Chisq diff** | **Df diff** | **Pr(>Chisq)** |
| --- | --- | --- | --- | --- | --- | --- | --- |
| fitBLCS_FA | 28 | 66202.68 | 66601.76 | 272.4489 | NA | NA | NA |
| fitBLCS_FA_corr_1_ | 29 | 66200.77 | 66592.72 | 272.5358 | 0.0884543 | 1 | 0.7661514 |

|  | **Df** | **AIC** | **BIC** | **Chisq** | **Chisq diff** | **Df diff** | **Pr(>Chisq)** |
| --- | --- | --- | --- | --- | --- | --- | --- |
| fitBLCS_FA | 28 | 66202.68 | 66601.76 | 272.4489 | NA | NA | NA |
| fitBLCS_FA_corr_2_ | 29 | 66201.92 | 66593.87 | 273.6875 | 1.248323 | 1 | 0.2638731 |
